# Correlates of protection for booster doses of the BNT162b2 vaccine

**DOI:** 10.1101/2022.07.16.22277626

**Authors:** Tomer Hertz, Shlomia Levy, Daniel Ostrovsky, Hannah Oppenheimer, Shosh Zismanov, Alona Kuzmina, Lilach M Friedman, Sanja Trifkovic, David Brice, Lin Chun-Yang, Yonat Shemer-Avni, Merav Cohen-Lahav, Doron Amichay, Ayelet Keren-Naus, Olga Voloshin, Gabriel Weber, Ronza Najjar-Debbiny, Bibiana Chazan, Maureen A. McGargill, Richard Webby, Michal Chowers, Lena Novack, Victor Novack, Ran Taube, Lior Nesher, Orly Weinstein

## Abstract

Variants of concern (VOC) of SARS-CoV2 and waning immunity pose a serious global problem. Overall, vaccination and prior infection appear to provide significant protection to the majority of individuals, but some remain susceptible to infection and severe disease. Rigorously identifying a broad spectrum of correlates of protection (COP) is necessary to identify these susceptible populations. The extent to which additional booster doses provide protection is also poorly understood. To address this need, we conducted a multicenter prospective study assessing the association between serological profiles and the risk for SARS-CoV-2 infection, comparing those vaccinated with three to four doses of Pfizer BNT162b2 vaccine. Of 608 healthy adults, 365 received three doses and 243 received four doses. During the first 90 days of followup, 239 (39%) were infected, of whom 165/365 (45%) received 3 doses and 74/243 (30%) four doses. We found that the fourth dose elicited a significant rise in antibody binding and neutralizing titers against multiple variants, and reduced the risk of symptomatic infection by 37% [95% I, 15% - 54%]. We identified several parameters based on IgG and IgA binding that were COPs. The strongest association with infection risk was reduced IgG levels to RBD mutants and IgA levels to VOCs, which was a COP in the three-dose group (HR=6.34, p=0.008) and in the four-dose group (HR=8.14, p=0.018). A combination of two commercially available ELISA assays were also associated with protection in both groups (HR = 1.84, p = 0.002; HR = 2.01, p = 0.025, respectively). Most importantly, we identified a subset of individuals with low antibody levels after three doses of vaccine that responded with a significant boost in neutralizing antibody titers after a fourth dose, but were still at significantly increased susceptibility to infection when compared to those who had pre-existing high levels of neutralizing antibodies. Thus, we identify a highly susceptible population that remains susceptible despite apparent responsiveness to vaccines. Further, we develop several specific and sensitive COPs that show dramatic effect sizes and may be utilized to identify individuals most at risk from future exposures.

---

Covid-19 is a disease caused by the SARS-CoV-2 virus and, for the past two and a half years, has been driving a worldwide pandemic. The pandemic has a broad spectrum of effects ranging from increased patient morbidity and mortality to impacting the global economy ^1^. The primary determinant in reducing this impact has been the rapid development of vaccines. The mRNA vaccines minimized infectiveness and reduced hospitalizations, severe disease, and death.^2^ However, not enough is known regarding the duration of protection or the schedule of boosting required ^3^ The SARS-CoV-2 virus has rapidly evolved, and variants of concern (VOCs) have swept the world every few months, with the omicron variant and its subvariants currently being the most common VOC.

Initially, the Pfizer-BioNTech mRNA vaccine was administered in two doses 21 days apart. However, the third dose of Pfizer-BioNTech was approved in August 2021 (6 months following the initial doses) in Israel and subsequently worldwide to combat the Delta variant and waning of vaccine-elicited antibody responses. Multiple studies reported that the third dose was very effective at inducing high neutralizing antibody levels^4,5^ and preventing complications of disease development and hospitalization ^6^. Towards the end of 2021, the Omicron BA.1 variant rapidly spread worldwide. BA.1 harbors up to 59 mutations throughout its genome, with 32 positioned within the spike and 15 within the receptor binding domain (RBD) ^7-9^. On January 2, 2022, the Israeli health ministry recommended the fourth dose of the Pfizer-BioNTech mRNA vaccine for immunocompromised groups, and a fourth dose was also offered to healthcare providers (HCPs) and people older than 60 years ^10^. Epidemiological studies on the fourth dose out of Israel demonstrated its effectiveness in reducing infection rates; however, these studies evaluated persons over 60 years old ^11,12^ or with a median age of 60 ^13^.

Here we report an interim analysis of the Clalit HCPs Booster study, which is a multicenter prospective trial in healthcare providers with increased risk of SARS-CoV-2 infection designed to identify novel correlates of protection (COP) for booster doses of the Pfizer-BioNTech vaccine. All participants were previously immunized with three doses of the Pfizer-BioNTech vaccine and without any documented history of SARS-CoV-2 infection. Participants were offered to receive the fourth dose and are currently longitudinally followed over six months. Blood samples were collected every 30 days, and participants with symptomatic SARS-CoV-2 infections were confirmed by PCR. Given the high attack rate of the Omicron BA.1 VOC in Israel, our study was powered to assess both individual baseline markers and pairwise combinations of these markers as COP. In line with previous reports, we demonstrate that the fourth dose induced significant binding and neutralizing antibody responses, and reduced infection rates. Importantly, we identify and validate several binding antibody COPs including those derived from commercial assays in clinical use. These assays are widely available and can identify individuals with increased risk of SARS-CoV-2 infection that may benefit from passive immunization and have clinical and epidemiological implications.

## Results

We enrolled 639 HCP from four medical centers between January 6 to February 9, 2022. Of the 639 enrolled participants, 31 were excluded (see supplement for details), and 608 individuals were included in the final analysis. Within our study, 243/608 individuals (40%) were vaccinated with the fourth dose of which 74 (30%) became infected, 365 (60%) received only three doses and 165 (45%) were infected (**Table 1**). In the current analysis, we analyzed immune responses in blood samples collected at enrollment and day 30, and infections during the first 90 days of follow-up (**Table 1**). The median number of days from the third vaccination to enrollment was 147. We analyzed outcomes at two time points: 30 days post enrollment and 90 days from study start date. Overall, 243 (39%) were vaccinated with the fourth dose. The median followup time was 76 days (IQR 75–77) for the four-dose group and 75 days (IQR 70– 77) for the three-dose group. The baseline characteristics of the participants within each vaccination group are shown in **Table 1**.

### Vaccination with the fourth dose elicited binding and neutralizing antibodies against multiple SARS-CoV-2 variants

We measured the magnitude of IgA and IgG antibodies binding to multiple SARS-CoV-2 spike antigens using an antigen microarray at day 0 and day 30 (**Fig. 1A-B, table S1, fig. S1**). We found that day 30 magnitudes of individuals receiving the fourth dose and were not infected by day 30 (n=127) were significantly higher than baseline for the Wuhan strain for both IgG (p<0.001) and IgA (p=0.004), as well as for IgA to SARS-CoV-2 variants (p<0.001). In contrast, IgG, but not IgA antibodies specific for the Wuhan strain (p<0.001) waned in individuals that did not receive a fourth vaccination (n=85), while both IgG and IgA reactive against the lSARS-CoV-2 variants (p<0.001), decreased in this group at the day 30 timepoint compared to enrollment. In addition, the decay of IgG binding to variants at day 30 was more pronounced than the decay of IgA responses (**Fig. 1B**).

**Fig. 1:**
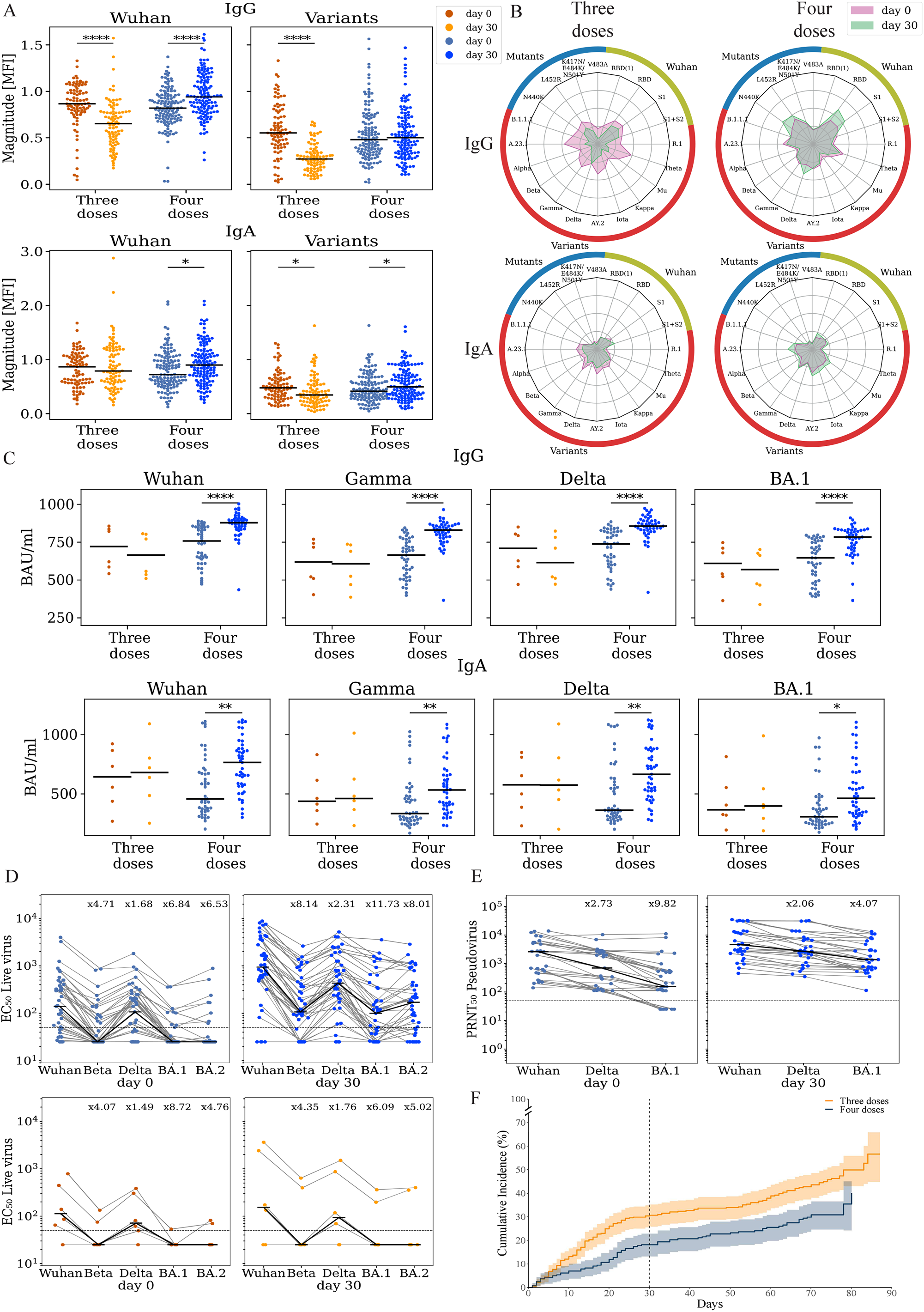
Vaccination with the 4th dose elicited binding and neutralizing antibodies against SARS-CoV-2. Responses of uninfected participants were analyzed at enrollment (day 0) and at day 30 using multiple serological assays (**A**) IgG and IgA magnitude to antigens from the Wuhan strain and SARS-COV-2 variants. Antigen microarrays spotted with RBD, S1 and spike proteins of the Wuhan vaccine strain and multiple other variants of concern were used to measure the magnitude of responses at day 0 (enrollment) and day 30 post enrollment. (**B**) Spider plots depicting the enrollment (pink) and day 30 (green) antibody levels to Wuhan antigens (green), variants of concern (red) and RBD mutants (blue). The average normalized magnitude to each antigen is plotted in individuals that received 3 or 4 doses. (**C**) IgG and IgA anti RBD ELISA binding titers for a subset of 74 participants (**D**) Live-virus neutralization EC50 titers of the same individuals in panel C. (**E**) Pseudovirus neutralization titers of uninfected individuals that received 4 (n=30, blue). (**F**) Cumulative incidence of SARS-CoV-2 infections in participants receiving three (n=365) vs. four doses (n=243) of the Pfizer vaccine. Four doses of the vaccine significantly reduced infection rates at day +30 (HR=0.55, p=0.002) and across all interim followup time (HR=0.63, p=0.003) as compared to three doses. * p < 0.05; ** p < 0.001; *** p < 0.0001; **** p < 0.00001.

To further assess the vaccine-elicited antibody responses, we selected a subset of 74 individuals for further in-depth immunogenicity assessment. Participants were selected based on their binding antibody levels to the RBD and S1 antigens of the Wuhan (vaccine) strain at enrollment. Of these, 58 individuals received the fourth dose, and 23 were infected within the first 30 days (**table S2**). We found that uninfected fourth dose individuals generated a significant rise in IgG and IgA titer against all four SARS-CoV-2 isolates measured by an ELISA against RBD of multiple variants (Fig. **1C** p <0.001). No significant rises were observed in individuals who only received three doses.

We next characterized the neutralization titers of these 58 participants using live-virus neutralization assays against Wuhan, Beta, Delta BA.1, and BA.2 (Fig. **1D**). In line with previous studies, we found a significant reduction in EC50 neutralization titers to BA.1 and BA.2. At (baseline fold drop of x6.8 and x6.5 respectively, post-vaccination fold drop of x11.7 and x8 respectively). The median baseline neutralization titer in both the three and four dose groups was below the estimated protective threshold (EC < 50, **(Fig. 1D**). We found a significant rise in the median neutralization titer of four-dose individuals at day 30 across all isolates (Fig. **1D** p < 0.001). However, 17 participants (29%) failed to generate any measurable rise in neutralization titer to BA.1 on day 30. We also characterized 47 participants from the immunogenicity subset using pseudovirus neutralization assays against the Wuhan, Delta, and BA.1 strains. We found a significant rise in titers at day 30 following the fourth dose, across all three isolates (Fig. **1D**). In contrast to the live-virus neutralization assay, all of the 30 four-dose and uninfected participants had measurable neutralization titers against BA.1 at day 30.

We then compared the IgG and IgA antibody responses of breakthrough infections in individuals that were infected within the first 30 days from enrollment (**fig. S2**). We found that overall infected participants had a significant rise in antibody levels regardless of whether they received a fourth dose of the vaccine or not. Moreover, there were no significant differences between day 30 titers of infected participants with 3 and 4 doses of the vaccine (**fig. S2**).

### A fourth dose of the Pfizer-BioNTech mRNA vaccine significantly reduced the risk of symptomatic SARS-CoV-2 infection

To assess the effect of the fourth dose on symptomatic SARS-CoV-2 infection, we estimated vaccine efficacy (VE) using a Cox model adjusted for age, occupation, medical center, and time from the third vaccination. We found that the VE at day 30 was 45.5% [95% CI, 19% - 63%], and the VE at the interim time point was 37% [95% CI, 15% - 53%] (**table S3-4**). Similar estimates were also obtained from a Poisson regression model adjusted for the proportion of daily positive PCR tests in Israel (**table S5-6**). Cumulative incidence curves of SARS-CoV-2 infection in the fourth dose group vs. the three dose group are shown in **(Fig. 1F**)

### Baseline binding antibody markers are associated with neutralizing antibody titers

The immunogenicity subset of 74 individuals included 38 individuals with low-baseline and 36 individuals with high-baseline IgG and IgA immune history to the S1 and RBD proteins of the Wuhan strain (see methods). (**Fig. 2A-B**, and **table S2**). Within this set, 58/74 (78.4%) received the fourth dose and 23/74 (31%) were infected by day 30 (**table S2**). We hypothesized that individuals from the low-baseline group would have significantly lower baseline neutralization titers. We found significant differences in the neutralization titer of the low-baseline and high-baseline groups at day 0 for the Wuhan, Beta, Delta and Omicron BA.1 strains (Fig. **2C**). Notably, 20% of the individuals in the low group had no detectable neutralization titers to the Wuhan strain, and none of them had any detectable titers to the Omicron BA.1 strain (**Fig. 2C**). In the high-baseline group, 14 individuals had no detectable titers to the Omicron BA.1 strain (**Fig. 2C**). However, post-vaccination with the fourth dose, neutralization titers were not significantly higher in the high-baseline group than in the low-baseline group (Fig. **2C**). In line with our overall observation of significant waning of individuals in the three dose group, we found out that ranking individuals who were un-infected by day 30 by both IgG or IgA responses to VOCs, IgG antibody responses waned more significantly in the high-baseline group vs. the low-baseline group (IgG p<0.001; IgA p<0.001, **(Fig. 2D)**. Interestingly, IgA responses waned significantly less than IgG responses, especially in the mid and high baseline groups (Fig. **2D**).

**Fig. 2:**
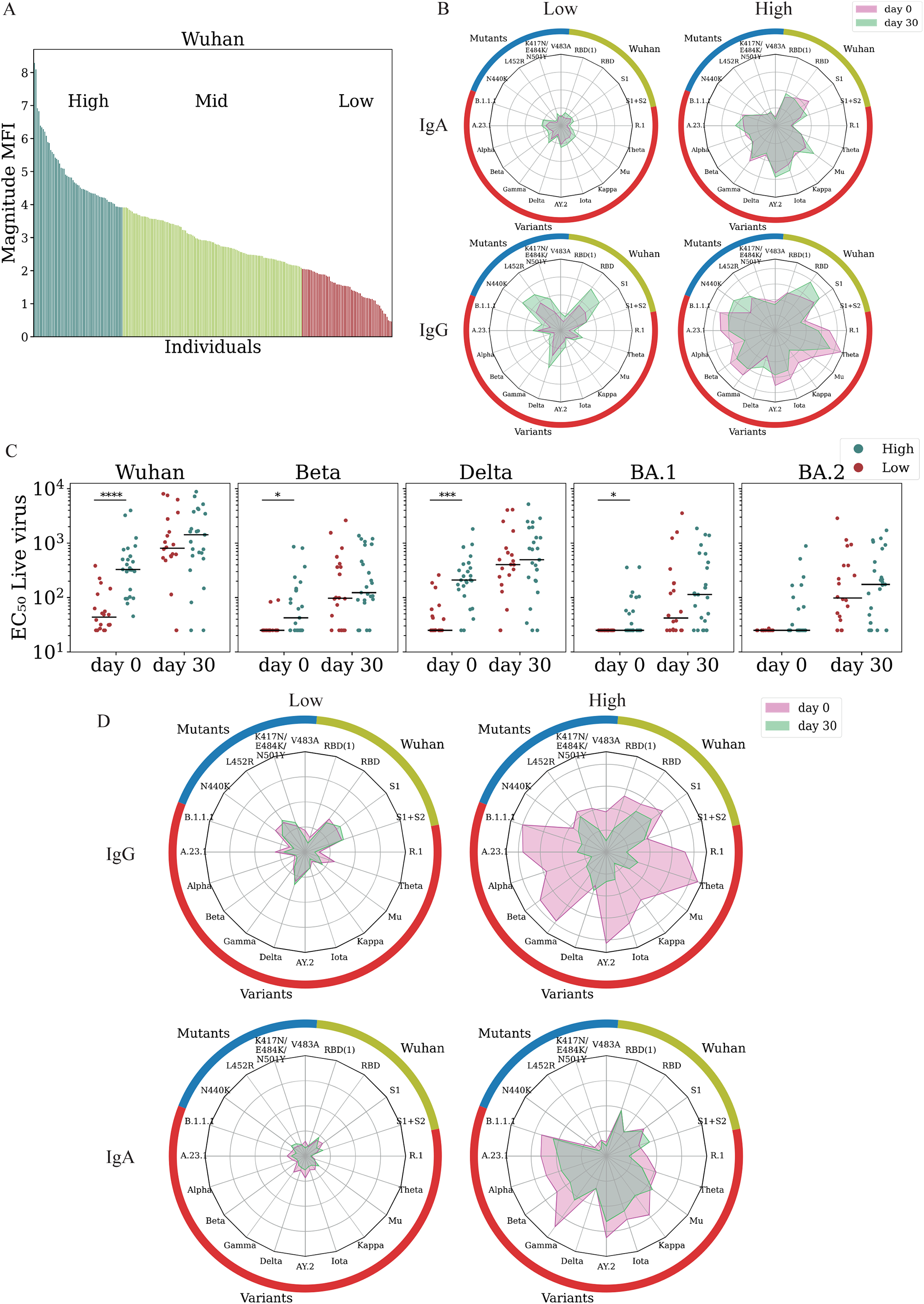
Ranking individuals using baseline binding antibody markers is associated with baseline neutralizing titers. **(A**) Ranking of 242 vaccinated individuals by their magnitude to SARS-CoV-2 Wuhan at enrollment. Each bar represents the magnitude of a single participant defined as the average response to the set Wuhan antigens (see **table S1**). Participants were divided into low (lowest quartile); mid (quartiles 2 + 3); and high (highest quartile) based on magnitude of IgA responses to Wuhan. (B) I**gA and IgG Antibody levels to SARS-CoV-2 antigens are not correlated**. Spider plots of the average normalized responses for a set of Wuhan antigens, RBD mutants and SARS-CoV-2 variants of concern. Responses of the low and high response groups are plotted separately for IgA (top) and IgG (bottom). Correlations between the IgG and IgA responses across the cohort were moderate (r < 0.310). (**C**) Live-virus neutralization titers of 58 vaccinated uninfected individuals at day 0 and day 30 from the low-baseline (red) and high-baseline (green) groups. (**D**) IgA and IgG Spider plots of 85 individuals that received 3 doses of the vaccine at day 0 (pink) and day 30 (green) comparing average day 0 (pink) and day 30 (green). Individuals were sorted by baseline response to SARS-CoV-2 VOCs. * p < 0.05; ** p < 0.001; *** p < 0.0001; **** p < 0.00001.

### Baseline binding IgA and IgG responses are correlates of protection for the Pfizer-BioNTech mRNA vaccine

We hypothesized that individuals with a low-baseline immune history to SARS-CoV-2 might be at an increased risk for SARS-CoV-2 infection. We found that in the three and four dose groups the IgA responses against the Wuhan RBD at enrollment were significantly higher in uninfected individuals as compared to infected individuals (p=0.042 and p=0.042, **Fig. 3A**). IgG responses against the RBD were not significantly associated with infection status (four doses: p=0.083; three doses p=0.281). IgA responses to the S1 protein were significantly higher in uninfected individuals as compared to infected individuals in the three-dose group (p=0.032, **Fig. 3A**) and responses to VOCs were higher in uninfected individuals in the four-dose group(p=0.048 **Fig. 3A**).

**Fig. 3:**
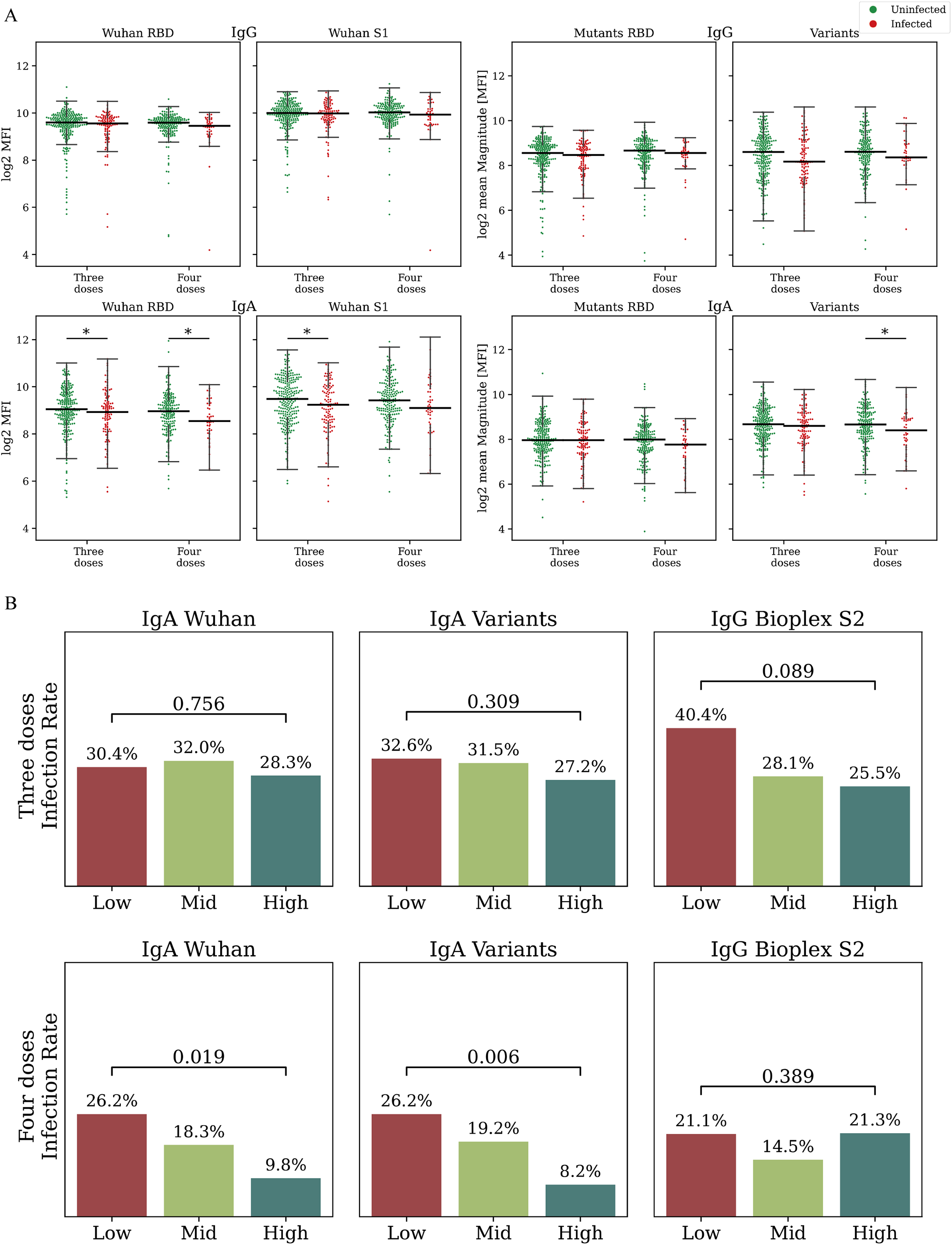
identifying baseline correlates of protection following three or four doses of the Pfizer-BioNTech vaccine. **(A)** Antibody levels of uninfected (green) and infected (red) individuals against Wuhan, RBD mutants and VOCs measured at enrollment. (**B**) Day 30 infection rates in low- mid- and high-baseline response groups ranked by baseline binding antibodies. Comparisons were conducted for three dose (top) and four dose (bottom) recipients separately. Individuals were ranked by IgA magnitude to Wuhan (Left), IgA to SARS-CoV-2 variants (Center), and by IgG S2 Bioplex (Right) (**D**) P-values were computed using a cox proportional hazard model, adjusted for age, occupation, medical center, and time from the third vaccination. * p < 0.05; ** p < 0.001; *** p < 0.0001; **** p < 0.00001.

To analyze baseline serological markers as COPs our primary analysis focused on the following antibody binding measures: (1) Wuhan magnitude - average response to the RBD, S1, and full-length S protein of the Wuhan strain; (2) Variants magnitude - average response to multiple SARS-CoV-2 variants of concern ; (3) RBD mutants magnitude - average response to multiple RBD mutants (**table S1**); (4) Wuhan S2 IgG levels (RAD Bioplex); (5) Wuhan IgG levels (Abbott Alinity). Magnitude baseline markers were computed for both IgG and IgA separately. Individuals were ranked and divided into quartiles, defining low, mid and high response groups. (**Fig. 2A**). We found that baseline IgG antibody levels of individuals within the low-mid- and high-baseline IgA groups were not significantly different from one another (Fig. **2B** and **fig. S3**). To quantify this further we computed the correlations between individuals’IgG and IgA magnitudes at baseline. We found that magnitudes were only moderately correlated one to another (r <0.310, **fig. S4**)

We compared the infection rates of the two groups at two time points: 30 days post-vaccination, and at the interim analysis timepoint which included 60-90 days of followup for all participants. We used a Cox regression model to compute the hazard ratios by comparing the low-baseline to the high-baseline groups. At the 30-day time point, IgA magnitude to the Wuhan strain was associated with infection risk in fourth dose recipients (HR=3.19, p = 0.019, **Fig. 3B, Fig. 4A, table S7**). IgA magnitude to SARS-CoV-2 VOCs was more strongly associated with infection risk (HR=4.45, p = 0.006, **Fig. 3B, Fig. 4B**). None of the IgG baseline markers were significantly associated with infection risk in the 4th dose vaccine group. However, in the three dose group, the Abbott Alinity was associated with infection risk (HR=1.59 p = 0.02, **Fig. 3B, Fig. 4C**). We then evaluated infection risk at the 60-90 day timepoint (**Fig. 4D, table S8**). At the 60-90 day follow-up time point, we found that in fourth dose recipients all markers were associated with infection risk (IgA Wuhan: HR=2.05, p = 0.041; S2: HR = 1.77, p = 0.025; Alinity: HR=1.65 p = 0.049, (**Fig. 4**). Within the third dose group the Bioplex S2 and Alinity were significantly associated with infection risk (S2: HR = 1.45, p = 0.022; Alinity: 1.54, p = 0.008; **Fig. 4)**. In our secondary analysis, we considered additional individual antigens as baseline correlates, and identified several additional baseline markers associated with infection status in both groups (**table S 9-10**).

**Fig. 4:**
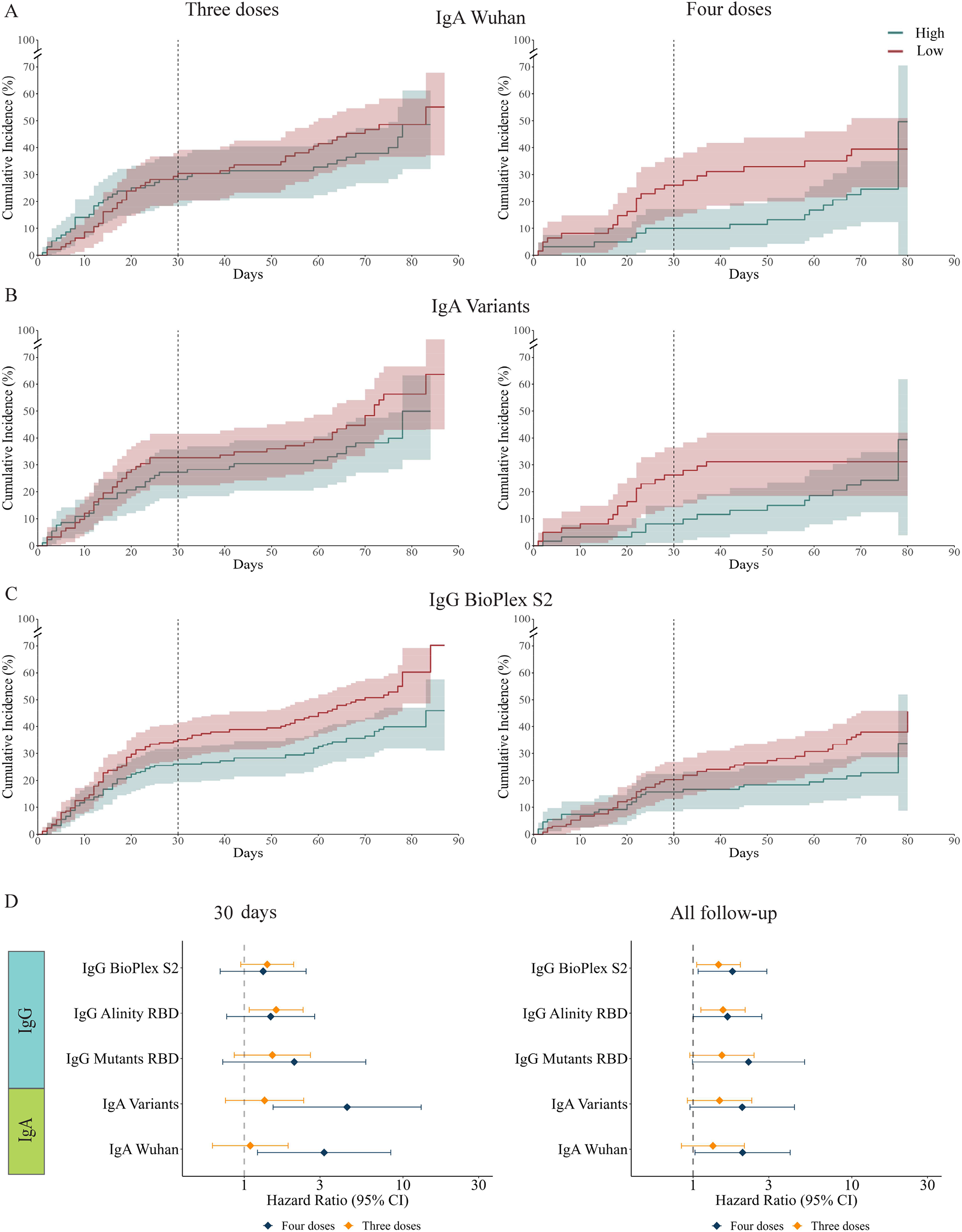
(A-C) Cumulative incidence plots of individuals in the low and high-baseline response groups as measured using: **(A)** IgA response to Wuhan, **(B)** IgA response to SARS-CoV-2 VOCs **(C)** IgG response to the S2 protein (RAD bioplex assay) **(D)** Hazard ratios for the five primary baseline markers comparing low to high baseline response groups for individuals vaccinated with three doses (orange) or four doses (blue) at day 30 (left) and the interim followup time point (right). Error bars denote the 95% confidence intervals. Hazard ratios were computed using a cox proportional hazard model adjusted for age, occupation, medical center, and time from the third vaccination.

### Combinations of IgG and IgA baseline markers as correlates of protection

Given the moderate correlations between IgG and IgA magnitudes (**fig. S4)**, we reasoned that combinations of IgG and IgA markers might provide improved COP against symptomatic SARS-CoV-2 infection. For each pairwise combination of baseline markers, we intersected the low-baseline and high-baseline groups and compared the infection rates of these groups. We found that combinations of baseline markers improved COPs in both groups, as compared to single baseline markers (**Fig. 5**, **table S 11-12**). For example, at the 60-90 day followup timepoint the infection rate in the low-baseline group ranked by IgG levels to RBD mutants, and IgA levels to VOCs, was 42.1% and only 13% in the high-baseline group in the fourth dose group (HR = 8.18, p = 0.018, **(Fig. 5A**). The same marker was also associated within the third dose group (HR = 6.34, p = 0.008, **(Fig. 5A**). Multiple marker combinations were significantly associated with infection status in both groups (Fig. **5C**). The combination of baseline IgA responses to the Wuhan strain and Iga responses to VOCs were significantly associated with infection risk at both timepoints, however, the association was stronger at the day 30 timepoint in fourth dose recipients (day 30 HR = 5.73, p = 0.009, interim timepoint: HR = 2.34, p = 0.051, **(Fig. 5C**). The combination of IgG Alinity RBD and IgG Bioplex S2 assays was also associated with infection status at the two time points for the third dose group and interim time point for the fourth dose group (**Fig. 5C**). Similar estimates for single markers and their pairwise combinations were obtained using a Poisson regression model as previously described **(table S 13-16)**.

**Fig. 5:**
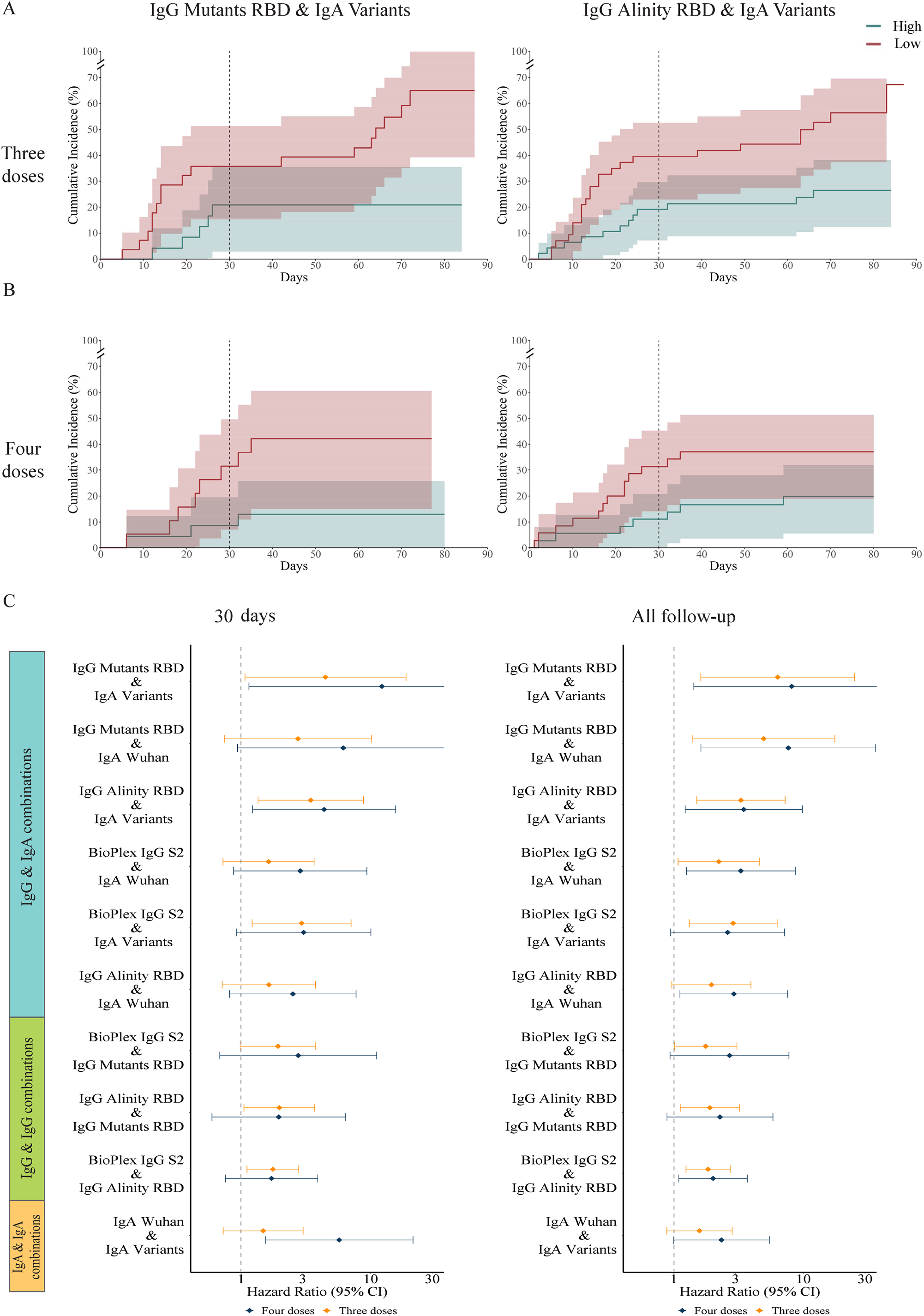
Combinations of IgG and IgA baseline markers are improved baseline correlates of protection. Pairs of baseline markers were used for ranking individuals using the intersection between the low and high groups of each baseline marker separately. Comparisons were conducted for 3rd and 4th dose recipients separately.(**A-B**) Cumulative incidence plots of individuals in the low and high-baseline response groups as measured using: (**A**) IgG RBD mutants and IgA SARS-CoV-2 VOCs (four doses n=42, three doses n=52); and (**B**) IgG Alinity and IgA SARS-CoV-2 VOCs (four doses n=71, three doses n=90). (**C**) Hazard ratios comparing low to high baseline response groups using pairwise combinations of baseline binding antibody markers for individuals vaccinated with three doses (orange) or four doses (blue). Error bars denote the 95% confidence intervals. Hazard ratios were computed using a cox proportional hazard model adjusted for age, occupation, medical center, and time from the third vaccination.

## Discussion

The main goal of this study was to assess the efficacy of the fourth vaccine and further identify novel binding antibody COPs against symptomatic SARS-CoV-2 infection during the omicron wave in vaccinated healthy individuals following one or two booster doses. Following the fourth dose, we showed that antibody levels were significantly elevated, which correlated well with neutralization titer against multiple SARS CoV-2 variants and overall infection protection. In addition, we found that IgA baseline markers against RBD mutants and spike VOCs are strongly associated with protection.

In our primary analysis, we used a set of five baselines IgA and IgG binding antibody markers, two of which are commercially available IgG assays. Using a simple quartile approach to define the low- and high-baseline response groups, we showed that infection rates in the low-response group were significantly higher than in the high-response group for both third and fourth dose recipients. Due to the moderate correlation between IgG and IgA baseline antibody levels of these five markers, we next considered pairwise combinations of these five baseline markers, including an IgG and IgA marker. By ranking individuals using such pairs of markers, the differences in infection rates between the low-baseline and high-baseline groups were more pronounced than single markers. This suggests that both IgA and IgG antibodies may play a protective role in preventing SARS-CoV-2 symptomatic infection. We also found that the combinations of IgA to Wuhan and variants were associated with infection risk in the fourth dose group at both time points and that all different combinations of IgG pairs were associated with infection status at the interim followup time point. Of particular interest is the combination of the Alinity RBD IgG and Rad Bioplex S2 IgG assays, both clinically approved assays widely used in clinical virology labs, and were COPs in both groups.

Studies on COPs for the Pfizer-BioNTech vaccine found that neutralizing antibody titers were a correlate of protection following two doses of the Pfizer-BioNTech vaccine^14–17^ and this was observed as well for other vaccines^18–21^. To the best of our knowledge, this is the first study to report COPs for booster doses of the Pfizer vaccine, and the first study to report COPs against infection with Omicron B.A1 ^15^. The COPs reported here were based on baseline binding antibody levels and not on neutralizing antibody titers. Binding antibodies measured by ELISA IgG antibody titers have been previously shown to be COPs for influenza ^22^and for SARS-CoV-2 ^23^. A clear advantage of binding antibody assays is that they can readily be measured at baseline for large cohorts, unlike neutralizing antibody assays.

IgA-based COPs have not been studied extensively, mainly due to the lack of standardized assays to quantify IgA levels in mucosal samples and their relatively low concentration in the blood. Burt et al. ^24^ found that both serum IgG & Mucosal IgA are important COPs against symptomatic influenza infection in a human challenge trial. They further showed that combinations of HAI titers and mucosal IgA titers were improved COPs against influenza infection. While IgA antibodies that offer protection from SARS-CoV-2 infection are primarily found in the mucosa, our reported IgA correlates were measured from the serum. A recent study in celiac patients reported that the gut mucosal and serological IgA repertoires share strong clonal overlap despite originating from different plasma cell compartments ^25^. Similar findings were reported for IgA plasmablasts from the serum and lungs following influenza vaccination ^26^. These studies suggest that while the serum IgA repertoire may not be directly involved in protection from infection of the respiratory tract, it may correctly reflect the mucosal IgA repertoire.

Our study measured the levels of a wide variety of antibodies against VOCs and against RBD mutants as COPs. Previous studies of binding antibody COPs were based on ELISA titers to a single viral variant, requiring one to choose a relevant variant. The approach used here utilizes a cross-reactivity score that integrates across all previous SARS-CoV-2 VOCs (excluding Omicron B.A1 or B.A2). The IgA and IgG magnitude to SARS-CoV-2 VOCs and to RBD mutants are measurements of the cross-reactive binding antibody responses. Due to the rapid evolution of the SARS-CoV-2 virus, it is difficult to identify the optimal single marker or variant which may best predict protection from infection for a novel VOC. Our data suggest that by using aggregate cross-reactivity measures to multiple variants, we can obtain more robust COPs even for strains that are antigenically distinct from previous strains, such as the Omicron B.A1 strain ^7^.

We show that ranking individuals using IgA & IgG markers was associated with significant differences in neutralizing antibody titers. Individuals with low-baseline binding antibodies had significantly lower neutralizing antibody titers than the individuals in the high-baseline group. Interestingly, while 35% of the individuals in the low-baseline group failed to develop detectable neutralizing antibody titers following a fourth dose, the majority of participants (65%) generated a significant rise in neutralizing antibody titers at day 30, demonstrating their ability to mount an adequate immune response following a fourth booster dose. In fact, on day 30, we found no significant differences between the individuals in the low-baseline and high-baseline groups, indicating that the fourth dose induced steeper rises in neutralizing antibody titers in the low-baseline group. These data suggest that most individuals in the low-baseline group may have low-baseline titers not due to a lack of ability to respond but possibly due to increased decay rates of their circulating anti-SARS-CoV-2 antibodies. Despite the significant rise in neutralizing antibody levels following the fourth dose, the number of infections in the low-baseline group (n=16) was significantly higher than in the high-baseline group (n=7, 43% vs. 20%, p=0.051).

We also demonstrated that a fourth dose of the vaccine generated a significant rise in both IgG and IgA binding antibodies, and neutralizing antibody titers using both live-virus and pseudovirus assays. At baseline, 59 (79.73%) of the 74 individuals from the immunogenicity subset had low detectable neutralizing titers to the Omicron B.A1 virus, and 56 (75.67%) had no titers to the Omicron B.A2 strain. Thirty days post-vaccination, only 13 (30%) of the 44 vaccinated uninfected individuals within the immunogenicity subset had no detectable titers to the B.A1 variant. These results are consistent with data reported by Regev-Yochai et al. ^13^ who reported a significant rise in neutralization titers 14 days post-vaccination with the fourth dose to both Delta and Omicron B.A1 variants. This rise in binding and neutralizing antibody titers was also associated with increased protection against symptomatic Omicron B.A1 infection. Our study also found that the antibody responses of individuals who only received three doses of the Pfizer-BioNTech vaccine continued to significantly wane over the first 30 days of the trial when individuals were at a median of 177 days from their third dose. This data is in agreement with a previous study that reported declines in binding and neutralizing titers up to 6 months post the second dose of the Pfizer-BioNTech vaccine ^27^. However, we found that IgG antibodies waned more significantly than IgA antibodies. We also found that individuals with hybrid immunity - i.e. that received 3 or 4 doses of the Pfizer-BioNTech vaccine and were subsequently infected with B.A1 had significant rises in neutralization titers to all five VOCs. A recent study reported that individuals with hybrid immunity had increased protection from Omicron infection as compared to vaccination alone^28^

One of the limitations of our study is that these findings are based on antibody magnitudes measured using an antigen array-based assay, which are currently not widely used in clinical settings. However, multiple other studies used this assay to profile SARS-CoV-2 antibody responses ^29–36^, and these arrays are now commercially available., Furthermore, we also found that the combination of IgG levels measured using the Alinity RBD assay and S2 levels measured using the Biorad Bioplex assay was a correlate of protection in both the third and fourth dose groups. While the association with infection risk was weaker for this combination as compared to combinations of IgG and IgA markers, these assays can be readily used by clinical labs to identify individuals with low baseline immune history against SARS-CoV-2 that are at an increased risk of infection.

In conclusion, our study demonstrated that combinations of IgA and IgG baseline antibody levels to SARS-CoV-2 VOCs are associated with protection from symptomatic infection. Importantly, our study identified a subpopulation of healthy adult individuals with low-baseline levels of IgA and IgG who are at increased risk for SARS-CoV-2 infection, despite receiving three or four doses of the Pfizer-BioNTech vaccine. These findings warrant further study into this group to assess if they are also at higher risk for severe disease or spread the infection more readily than others.

## Supporting information

supplements_Correlates_of_protection_for_booster_doses_of_the_BNT162b2_vaccine

## Data Availability

All data produced in the present study are available upon reasonable request to the authors

## Acknowledgments

We would like to acknowledge the following for their contribution to this study: Lihi Marciano, Ruthie Bechor, Hana Kahanov-Edelstein, Galit Carmon, Moran Raad, and Natalie Samiande.

## Author contributions

T.H., V.N., R.T., <u>L.NE</u>., O.W. designed the study; Y.A,S., G.W., B.C., M.C., contributed to the methodology; G.W, R.D.N., B.C., V.N. <u>L.NE</u>., were involved in clinical recruitment and clinical study; S.L., H.O., S.Z., A.K., L.M.F., S.T., D.B., L.Y.C., Y.A.S., O.V., M.A.M., R.W., R.T. carried out the experiments; D.A., M.L.C., N.K.A. provided laboratory support, D.O., N.L., N,V. provided statistical guidance; T.H., S.L., D.O., H.O., L.M.F.,S.Z., Y.A.S., L.N., V.N., R.T.,<u>L.NE</u>. were involved with analysis and interpretation. T.H., S.L., D.O., R.T., <u>L.NE</u>., drafted the manuscript; T.H., Y.A.S., M.A.M., R.W., M.C., R.T., <u>L.NE</u>. critically revised the manuscript; all authors approved the final version for publication

## Data and materials availability

All data needed to evaluate the conclusions in the paper are present in the paper or the supplementary materials. All analysis code will be available upon request.

