## supplements_Correlates_of_protection_for_booster_doses_of_the_BNT162b2_vaccine for "Correlates of protection for booster doses of the BNT162b2 vaccine"

### Abstract

**Introduction**: Variants of concern (VOC) of SARS-CoV2 and waning immunity pose a serious global problem. Herein, we aimed to identify novel correlates of protection (COPs) against symptomatic SARS-CoV-2 infection.

**Methods**: We conducted a Multicenter prospective study assessing the association between serological profiles and the risk for SARS-CoV-2 infection, comparing those vaccinated with three to four doses of Pfizer BNT162b2 vaccine.

**Results:** Of 608 healthy adults, 365 received three doses and 243 received four doses. During the first 90 days of followup, 239 (39%) were infected, of whom 165/365 (45%) received 3 doses and 74/243 (30%) four doses. We found that the fourth dose elicited a significant rise in antibody binding and neutralizing titers against multiple variants, and reduced the risk of symptomatic infection by 37% [95% I, 15% - 54%]. We found several binding IgG and IgA markers and their combinations that were COPs. The strongest association with infection risk was IgG levels to RBD mutants and IgA levels to VOCs, which was a COP in the three-dose group (HR=6.34, p=0.008) and in the four-dose group (HR=8.14, p=0.018). A combination of two commercially available ELISA assays were also associated with protection in both groups (HR = 1.84, p = 0.002; HR = 2.01, p = 0.025, respectively). In a subset, comparing those with low to high antibody levels before 4th dose, despite a significant rise in neutralizing antibody titers against both omicron variants, the number of infections in the low group (n=16) was significantly higher than in the high group (n=7, 43% vs. 20%, p=0.051).

**Conclusions:** We demonstrated that following immunization with three or four vaccine doses, combinations of IgA and IgG levels are associated with protection from symptomatic infection. In addition, we identified a subpopulation of healthy adult individuals with low-baseline levels of antibodies after 3 doses which are at an increased risk for SARS-CoV-2 infection despite receiving a fourth dose. These findings warrant further study of this group, assessing whether they are at a higher risk for developing severe disease or may spread infection more readily than others.

**Supplementary Materials**

Materials and Methods

Figs. S1 to S4

Tables S1 to S16

References (1–3)

### Methods

#### Study design and setting

This Clalit HCP Booster study is an ongoing prospective cohort study designed to assess the association between different serological profiles and risk for SARS-CoV-2 infection, comparing those vaccinated with three doses of Pfizer-BioNTech vaccine (Three-dose) to those who received a fourth booster dose (Four-dose). For this multicenter study, we enrolled HCPs at four medical centers managed by Clalit Health Services (CHS), the largest integrated payer-provider healthcare organization in Israel with 4.7 million members. The medical centers are spread across Israel: Ha'Emek and Carmel Medical Centers in northern Israel, Meir Medical Center in the central region, and Soroka University Medical Center in southern Israel.

We enrolled HCPs over 18 years of age immunized with three doses of the Pfizer-BioNTech vaccine; the last dose was given at least three months prior to enrollment. We excluded people with prior SARS-CoV-2 infection or a history of receiving chemotherapy or immunosuppression therapy within the last three months (including immunomodulatory drugs, biological agents, and any immunosuppressive drug).

Data related to all SARS-CoV-2 PCR tests in Israel is collected centrally by the Israeli Ministry of Health (MoH) and is updated daily into CHS's electronic medical records. We, therefore, collected data directly from the CHS database. Additionally, all participants completed a brief questionnaire at enrollment and at every monthly visit. Data collection and management for the study were conducted using REDCap (Vanderbilt University, Nashville, TN, USA).

Followup time was calculated in person-days. At the interim time point analyzed here, participants were followed for three months. The time to infection for individuals receiving three doses was measured from the day of enrollment and for individuals receiving four doses of vaccine, starting from the eighth day after receiving the fourth vaccine dose.

**Statistical analysis**

The results are presented as the mean (SD) for continuous variables and as the total patients (percentage of total patients) for categorical data. A t-test was used to compare the continuous variables and chi-square test for categorical data, using Fisher's exact test if needed. In addition, we used Mann–Whitney test to compare variables without normal distribution. In the primary analysis, we compared the rates of Covid-19 infection among different serological response groups. The Kaplan-Meier estimator was used to construct cumulative incidence curves describing the infection rate. We used the Cox proportional hazards model adjusted for age, occupation (physician/nurse or administrative/support staff), medical center, and time from the third vaccination to assess risk of infection. The risk was defined as the fold increase in the hazard of being infected. We used calendar time as the time scale to account for fluctuations in infection rates. We used the same model to assess vaccine efficacy (VE), defined as one minus the hazard ratio. Previous studies demonstrated that VE following a fourth dose of the Pfizer mRNA vaccine wanes after 30 days [1]. In line with these findings, we found that the cumulative incidence curves of the third and fourth dose recipients in our study became parallel around day 30 (Fig. **1F**), indicating similar infection rates from this time-point and on. Therefore, we estimated VE at day 30 and also analyzed VE for the entire interim followup time in which all participants were followed for at least 60 days. Since vaccination with a fourth dose may modify VE, we analyzed COP separately in the three and four-dose groups. Furthermore, due to the possible time-limited VE, we analyzed these differences at day 30 and at the interim followup time-point.

To explore the robustness of our estimates, we performed a sensitivity analysis for the main results. We applied a Poisson regression adjusted to the same variables aforementioned and the daily proportion of positive PCR tests. In addition, we added subject IDs as a random effect to account for repeated measures. This analysis defines risk as to the fold increase in the incidence rate ratio and VE as 1 minus the incidence rate ratio.

Analyses were performed using R software, version 4.1.2, and the additional freely available R software packages “data.table,” version 1.14.2, and “survminer,” version 0.4.9, as well as using Python version 3.10.

**Ethics**

The study was approved by the CHS Central Institutional Review Board of the Israeli ministry of health (0404-21-SOR-C). All participants provided written informed consent. The report follows the STROBE methodology [2].

**Live-virus neutralization assays**

Viruses in neutralization assays were isolated from de-identified, discarded nasal swabs and grown in VeroE6 cells ectopically expressing both TMPRSS2 and human ACE2 (VE6/T2/ACE2; provided by Dr. Barney Graham at VRC, NIAID, NIH). Briefly, 100uL of swab suspension was inoculated onto VE6/T2/ACE2 cells seeded in 6 well tissue culture plates and incubated at 37°C, 5% $CO_2$ until 90% cytopathic effect (CPE) was observed. The presence of the virus was confirmed by BD Veritor System for rapid detection of SARS-CoV-2 (Catalog # 256082). Virus stocks were subsequently expanded using a VeroE6 cell line ectopically expressing TMPRSS2 (VE6/T2; from JCRB Cell Bank, Japan (https://cellbank.nibiohn.go.jp/english/)). Briefly, VE6/T2 cells were inoculated with the virus at a 1:50 dilution and incubated at 37°C, 5% $CO_2$ until 90% CPE was observed. Virus stocks were tittered in VE6/T2 cells to determine a 50% tissue culture infectious dose (TCID50). Cells in a 96 well format were inoculated with a 1:10 serially diluted virus stock for 72 hours. Wells were stained with 0.1% crystal violet solution to visualize cells. Infectious dose titers were determined using the Reed and Muench method.

Live-virus microneutralization assays were performed in a 96-well format. Half-log serial dilutions of heat-inactivated plasma or sera (1 hour at 56°C), starting at a 1:50 dilution, were incubated with 250 $TCID_{50}$ of infectious SARS-CoV-2 virus at a 1:1 ratio for 1 hour at 37°C. The serum/plasma mixture was then added to VE6/T2 cells and incubated at 37°C, 5% $CO_2$ for 24 – 48 hours. Following incubation, cells were fixed with 4% formaldehyde for 30 mins, washed with PBS, and incubated with a block/permeabilization buffer (PBS supplemented with 3% Bovine Serum Albumin and 0.2% Triton-X-100) for 30 minutes. Rabbit anti-SARS CoV-2 NP mAb (Sino Biologicals Cat # 40143-R040) at a 1:2000 dilution was added for 1 hour and cells were washed with PBS supplemented with 0.5% Tween (PBST) before incubation with a secondary goat anti-rabbit IgG–HRP conjugated antibody (Cell Signaling Cat# 7074S)) at a 1:3000 dilution for 1 hour. Finally, cells were washed with PBST and incubated with TMB for 10 mins before 1N sulfuric acid (Fisher Scientific Cat #SA212-1) was added to stop the reaction. The optical density was measured at 450nm on a Biotek Synergy plate microplate reader. To compute EC50 values, we subtracted the mean of the negative control from all wells, and fitted values using a five parameter logistic regression model (5PL) using the python scipy package. The readout of the positive control was used as the maximal response for curve fitting.

**Pseudovirus neutralization assays**

Pseudotyped viruses were generated in HEK293T cells. Pseudoviruses were generated following transfection of , LTR-PGK luciferase lentivector into HEK293T cells together with lentiviral packaging plasmids coding for Gag, Pol Tat Rev, and the corresponding wild type or mutate SARS CoV-2 spike envelopes. Transfections were performed in a 10cm format and the supernatant containing virus was harvested 72hr post-transfection, filtered, and stored at -80°C - as previously described (Krasnopolsky et al., 2020). Pseudovirus quality control and titers were determined by transducing HEK293T cells expressing ACE2 (HEK-ACE2) that were plated in a 12-well plate. 24 h later, transduction was monitored serial dilutions of pseudovirus were used ts. 48 hr. post-transduction, cells were harvested and analyzed for their luciferase readouts. p24 ELISA measurements were also conducted to ensure equal loads.
Neutralization assays were performed in a 96 well format, in the presence of pseudotyped viruses that were incubated with increasing dilutions of the tested sera (1:50; 1:250: 1;1250: 1:6250; 1:31250) or without sera as a control. Virus and sera were incubated for 1hr. at 37°C, followed by transduction of HEK-ACE2 cells for an additional 12 hr. 72hr post-transduction, cells were harvested and analyzed for luciferase readouts according to the manufacturer protocol (Promega). Neutralization measurements were performed in triplicates using an automated Tecan liquid handler and readout was used to calculate $NT_{50}$ – 50% inhibitory titers concentration. All experiments were run in technical duplicates or triplicates.

**ELISA assay**

Recombinant SARS-CoV-2 proteins purchased from Sino Biological include the full-length spike protein (40589-V08H) and RBD (40592-V08H) from the Wuhan-Hu-1 isolate, the RBD of the B.1.617.2 (Delta) variant (40492-V08H90), and the RBD of the BA.1 (Omicron) variant (40592-V08H121). Expression plasmids for the nucleocapsid (N) protein from the Wuhan-Hu-1 isolate and the RBD of the B.1.1.28 or P.1 (Gamma) variant were obtained from Florian Krammer. Plasmids were transfected into Expi293F cells using an ExpiFectamine 293 transfection kit (Thermo Fisher Scientific, A14524) as previously described (Amanat, F. et al. PMID32398876). Supernatants from transfected cells were harvested and purified with a Ni-NTA column. The resulting purified proteins were used for ELISA analysis of serum samples.

For antibody detection by ELISA, 384-well microtiter plates were coated overnight at 4° with recombinant proteins diluted in PBS. Optimal concentrations for each protein and isotype were empirically determined to optimize sensitivity and specificity. The N protein was coated at 1 μg/ml for IgG detection and 2 μg/ml for IgA detection. The full-length spike protein was coated at 2 μg/ml for IgG detection and 4 μg/ml for IgA detection. All RBD proteins were coated at 4 μg/ml for IgG and IgA detection. The following day plates were washed three times with 0.1% PBS-T (0.1% Tween-20) and blocked with 3% OmniblokTM non-fat milk (AmericanBio; AB10109-01000) in PBS-T for one hour. Plates were washed three times with 0.1% PBS-T immediately before the addition of diluted samples. Prior to dilution, plasma or serum samples were incubated at $56^{\circ}$c for 15 minutes and then diluted in 1% milk in PBS-T. Diluted samples were added to the blocked plates and incubated for 90 minutes at room temperature. The plates were washed three times and incubated for 30 minutes at room temperature with secondary antibodies diluted in 1% milk in PBS-T: anti-IgG (1:10,000; Invitrogen, A18805) or anti-IgA (1:2,000; Southern Biotech, 2050-05). The plates were washed and incubated at room temperature with SIGMAFAST OPD (Sigma-Aldrich; P9187) for eight minutes. The chemiluminescence reaction was stopped by the addition of 3N HCl and absorbances were measured at 490 nm on a microplate reader. To control for plate-to-plate variability, the same positive and negative control samples are included on each plate. In addition, the WHO international standard from the National Institute of Biological Standards and Control (NIBSC, cat# 21/234) was included on each plate. The WHO standard contained 817, 832, and 713 binding antibody units (BAU)/ml for the RBD, full-length spike, and N IgG, respectively. For the IgA, we previously calculated the BAU/ml of our control samples using the NIBSC standard 20/136, which was 1000 BAU/ml for all antigens and isotypes. All OD values were converted to BAU/ml using the reference standards on each plate.

**Antigen microarray spotting.** Recombinant SARS-CoV-2 proteins were spotted onto *N*-hydroxysuccinimide ester–derivatized Hydrogel slides (H slides) using a Scienion Sx non-contact array spotter. The proteins were purchased from Sino Biological (China) or were obtained as gifts through BEI Resources (NIAID, NIH), from ACROBiosystems, as listed in **table S1**. Each recombinant SARS-CoV-2 protein was diluted in PBS to the concentration of 130 μg/ml and was spotted in 3 concentrations (65, 35, and 16.25 μg/ml) in 0.0025% Triton X-100. Spot volumes ranged between 300-360 pL. Each antigen at each concentration was spotted in triplicate. Sixteen identical microarrays were spotted on each microarray slide. All samples were profiled using microarrays from a single printing batch, which included 140 microarray slides each containing 16 arrays per slide.

**Antigen microarray assay.** Array slides were blocked with 4 ml chemical blocking solution per slide (50 mM ethanolamine, 50 mM borate, pH 9.0) for 1 hour at room temperature (RT) on a shaker. After blocking the liquid was vacuumed, the slide was washed 2 times for 3 minutes in a washing buffer (0.05% tween-20 in PBS), 2 times for 3 minutes in PBS and an additional

3 minutes wash in double deionized water (DDW). Every wash was with 3 ml of liquid per slide on a shaker at RT. Samples were diluted in a hybridization buffer (1% BSA / 0.025% tween-20 in PBS). Human serum samples were diluted at 1:1000 for IgG characterization and 1:100 for IgA characterization. Following 2 hours of incubation, the slides were dried by centrifugation at RT for 5 minutes at a speed of 2000 rpm in a slide holder padded with Kim wipes, loaded on divided incubation trays (PepperChips, PepperPrint, Germany), and then the samples were added and hybridized with the arrays for 2 hours at RT on a shaker. After hybridization, the samples were discarded and the slides were washed twice with a washing buffer and twice with PBS as described above. After washes, the slides were incubated for 45 minutes on the shaker at RT with a fluorescently labeled polyclonal secondary antibody in the hybridization buffer. The secondary antibody for IgG was Alexa Fluor® 647 affinipure Donkey Anti-Human IgG (H+L), cat# 709-605-149, Jackson ImmunoResearch at 1:1000 dilution. The secondary antibody for IgA was Alexa Fluor® 647 affinipure Goat Anti-Human serum IgA $\alpha$ Chain Specific, cat# 109-605-011, Jackson ImmunoResearch at 1:5000 dilution.To detect bound immunoglobulins, slides were scanned on a three-laser GenePix 4400 scanner. Images were analyzed using GenePix Pro version 7 to obtain the mean fluorescence intensity (MFI) of each spot after subtracting the mean local background fluorescence intensity ($0 \leq$MFI $\leq$ 65,000).

**Antigen microarray analysis.** The array results were uploaded to a python pandas dataframe and analyzed using python scripts. Since each antigen at each concentration was spotted in triplicate, the median fluorescent intensity (MFI) of each triplicate was calculated. During each experiment, a negative control array was hybridized with the hybridization buffer only. The background staining of the negative control array was subtracted from each other array. Since the antigens were spotted in serial concentrations, a 5-parameter logistic regression model was used to fit curves to the measured MFI across all antigen concentrations, and the area under the curve (AUC) was calculated for each antigen. The magnitude of antibody response to a group of antigens was defined as the sum of MFI AUC of all the proteins included in the group. We summed 3 groups of antigens for magnitude computation: 1. Wuhan - Wuhan spike and RBD antigens (whole S1+S1 protein, S1 subunit alone, RBD alone); 2. Variants - whole spike antigens (S+S2) of non-Wuhan SARS-CoV-2 variants: B.1.1.1, A.23.1, Alpha, Beta, Gamma, Delta, AY.2, Iota, Kappa, Mu, Theta and R.1; 3. RBD Mutants - Wuhan RBD sequence including specific mutation: V483A, K417N/E484K/N501Y, L452R, or N440K. We used mean centering [3] to normalize results across different experiments.

**Ranking participants by baseline markers.** We used various different baseline markers to rank participants from highest to lowest. First, rankings were computed within the three-dose and four-dose groups separately. Then, after ranking participants, we used a quartile analysis to define three baseline groups: (1) ‘low’ - lowest quartile; (2) ‘mid’ - quartiles 2 and 3; and (3) ‘high’ - highest quartile.

For the two commercially available serological assays tested, we also used a single threshold approach to divide participants into a low baseline and high-baseline group as follows: (1) BioPlex 2200 SARS-CoV-2 IgG panel, S2 (Biorad Laboratories, Cal, USA) - we used the clinical cutoff provided by the manufacturer (< 10) to define the ‘low’ group, and all individuals with a titer >= 10 were defined as ‘high’; (2) SARS-CoV-2 IgG II Quant RBD of S1 subunit (Alinity i, Abbott, USA): Since all participants had a measurable titer at baseline, we used the median titer (4560) as a single threshold. Participants with titers < 4560 were assigned to the low-baseline group and participants with titer >= 4560 were defined as the high-baseline group.

**Combining clinical markers.** To combine the rankings of two serological baseline markers, we considered the intersection of the two low-baseline and high-baseline groups to define the low and high groups of each combination. We only considered combinations of markers that were significantly associated with protection for either three-dose or four-dose individuals, considering both the 30-day and interim followup timepoints.

**Immunogenicity subset selection.** We used baseline responses of participants to rank them based on their responses to the S1 and RBD antigens of the Wuhan wildtype strain. We selected 40 participants with low baseline antibody levels (low-baseline) and 40 participants with high antibody levels (high-baseline) for an in-depth immunogenicity assessment at the baseline and day 30 post-vaccination time points (**table S2**). Participants were selected at enrollment and included 58 (42.92%) 4th dose recipients. 23 (17.02%) participants were infected within the first 30 days of the trial (**table S2**).

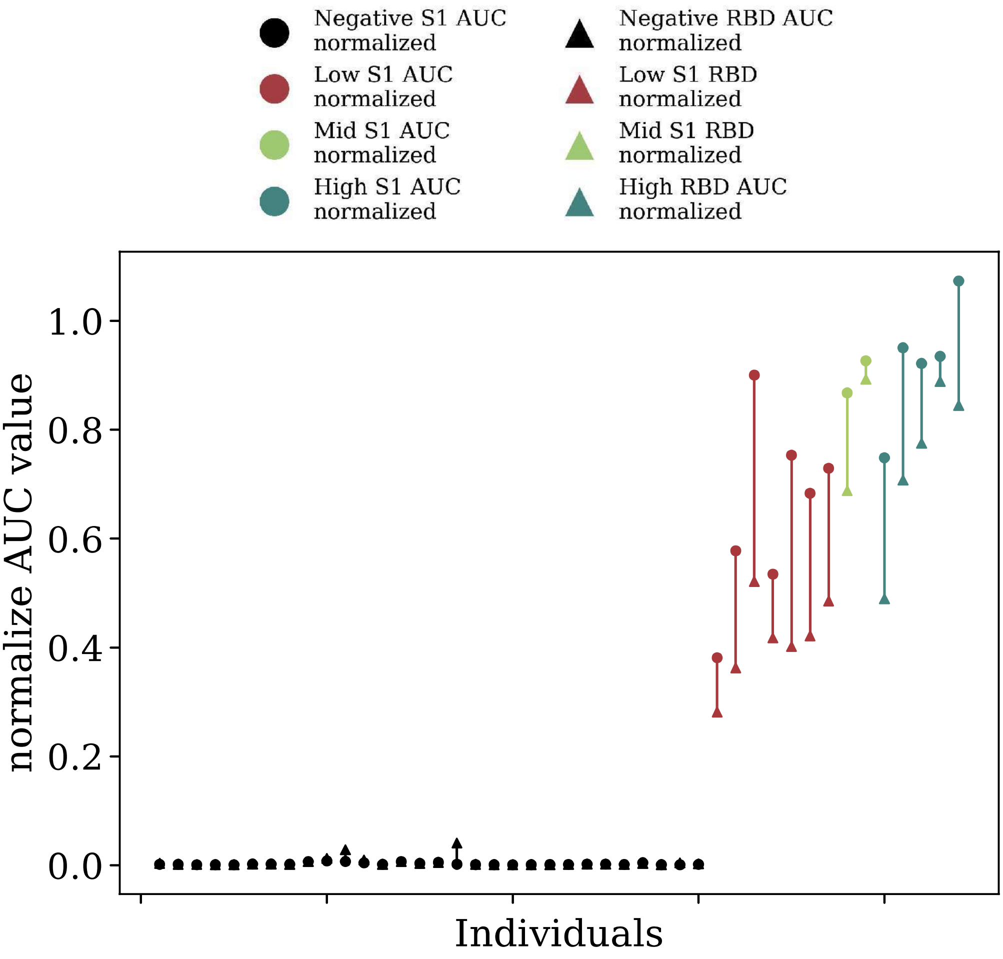
Negative S1 AUC normalized
Low S1 AUC normalized
Mid S1 AUC normalized
High S1 AUC normalized
Negative RBD AUC normalized
Low S1 RBD normalized
Mid S1 RBD normalized
High RBD AUC normalized
normalize AUC value
0.0
0.2
0.4
0.6
0.8
1.0
Individuals

**fig. S1** Negative control microarray assay

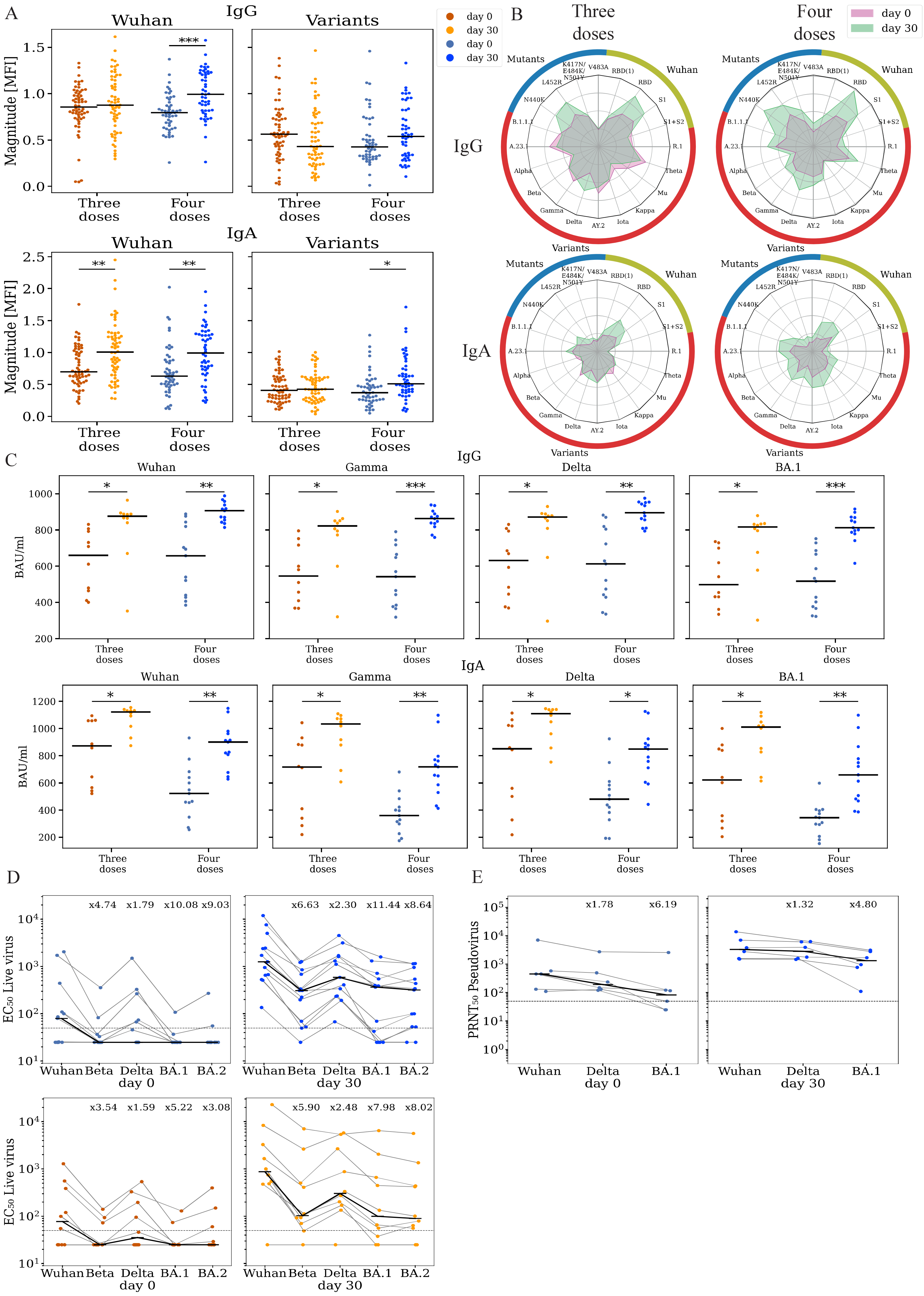
A
IgG
Wuhan
Variants
Magnitude [MFI]
Three doses
Four doses
***
IgA
**
*
day 0
day 30
B
Three doses
Four doses
Mutants
Wuhan
Variants
K417N/ E484K/ N501Y
V483A
RBD(1)
RBD
S1
S1+S2
R.1
Theta
Mu
Kappa
Iota
AY.2
Delta
Gamma
Beta
Alpha
A.23.1
B.1.1.1
N440K
L452R
C
Gamma
Delta
BA.1
BAU/ml
D
EC50 Live virus
x4.74 x1.79 x10.08 x9.03
x6.63 x2.30 x11.44 x8.64
x3.54 x1.59 x5.22 x3.08
x5.90 x2.48 x7.98 x8.02
Wuhan
Beta
BA.2
day 0
day 30
E
PRNT50 Pseudovirus
x1.78 x6.19
x1.32 x4.80

**fig. S2: Infection with omicron elicited binding and neutralizing antibodies against SARS-CoV-2.** Responses of individuals that were infected with omicron within the first 30 days after enrollment were analyzed at enrollment (day 0) and at day 30 using multiple serological assays. Individuals that received three or four doses of the vaccine were analyzed separately (**A**) IgG and IgA magnitude to antigens from the Wuhan strain and SARS-COV-2 variants. Antigen microarrays spotted with RBD, S1 and spike proteins of the Wuhan vaccine strain and multiple other variants of concern were used to measure the magnitude of responses at day 0 (enrollment) and day 30 post enrollment. (**B**) Spider plots depicting the enrollment (pink) and day 30 (green) antibody levels to Wuhan antigens (green) , variants of concern (red) and RBD mutants (blue). The average normalized magnitude to each antigen is plotted in individuals that received three or four doses. (**C**) IgG and IgA anti RBD ELISA binding titers for a subset of 74 participants (**D**) Live- virus neutralization EC50 titers of the same individuals in panel C. (**E**) Pseudovirus neutralization titers of uninfected individuals that received four doses (n=13, blue). (**F**) Cumulative incidence of SARS-CoV-2 infections in participants receiving three doses (n=365) vs. four doses (n=243) of the Pfizer vaccine. Four doses of the vaccine significantly reduced infection rates at day +30 (HR=0.55, p=0.002) and across all interim followup time (HR=0.63 , p=0.003) as compared to three doses. * $p < 0.05$; ** $p < 0.001$; *** $p < 0.0001$; **** $p < 0.00001$.

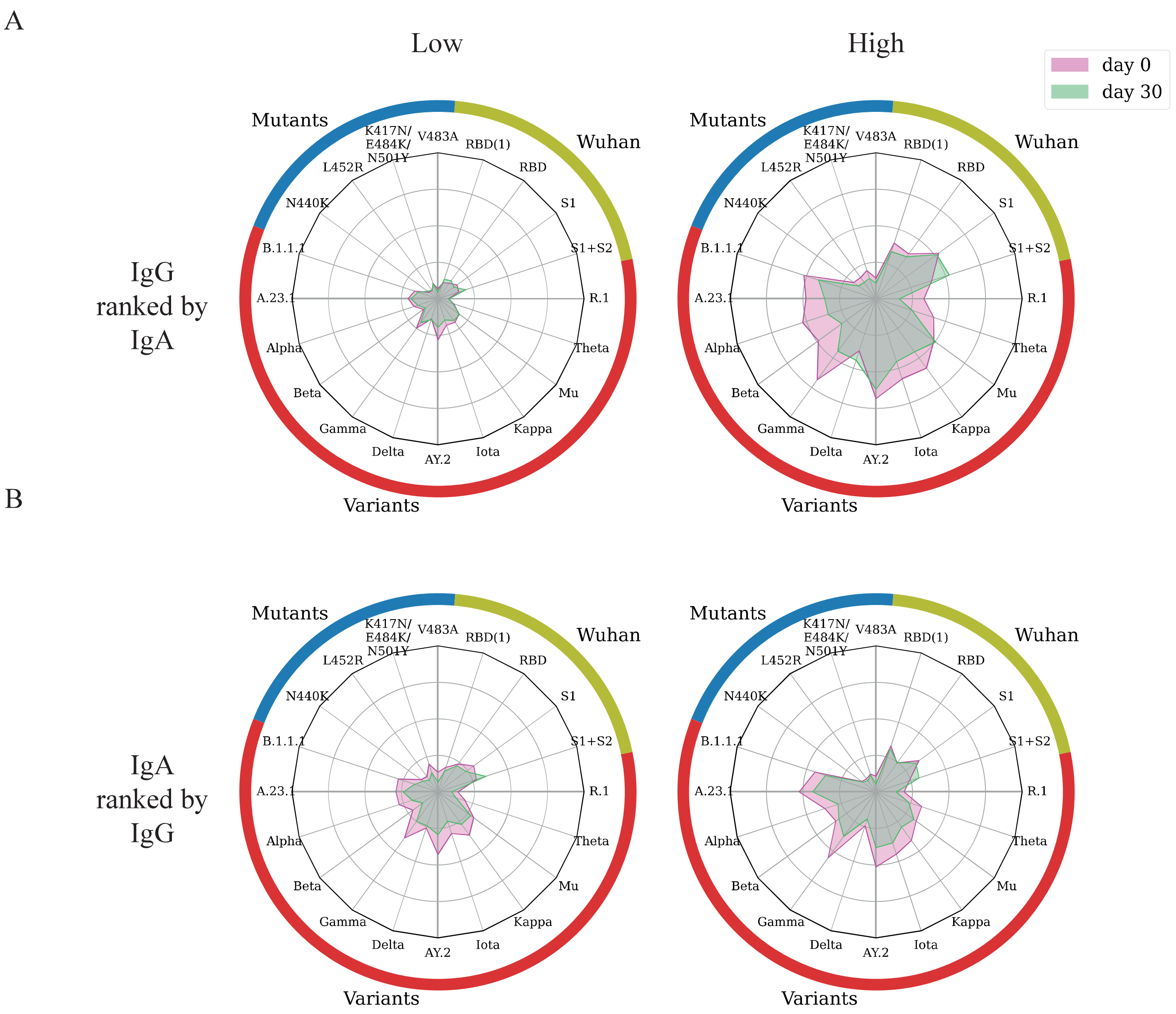
A
Low
High
day 0
day 30
IgG
ranked by
IgA
Mutants
Wuhan
Variants
K417N/
E484K/
N501Y
V483A
RBD(1)
RBD
S1
S1+S2
R.1
Theta
Mu
Kappa
Iota
AY.2
Delta
Gamma
Beta
Alpha
A.23.1
B.1.1.1
N440K
L452R
B
IgA
ranked by
IgG

**fig. S3: The decay of anti SARS-CoV-2 spike and RBD IgG antibodies is stronger than IgA antibodies**. We analyzed the antibody levels of 85 individuals that received three doses of the Pfizer vaccine and were not infected by SARS-CoV-2 up to day 30 (A) The 85 individuals were ranked according their IgA magnitudes to SARS-CoV-2. Data presented include the low-baseline (n=22) and high-baseline (n=22) groups. The IgG binding profiles of the low- and high-baseline IgA groups at day 0 (cyan) and day 30 (orange) are presented . (B) The 85 individuals were ranked according to their IgG magnitudes to SARS-CoV-2 VOCs. The IgA binding profiles of the low- and high-baseline IgG groups at day 0 (cyan) and day 30 (orange) are presented.

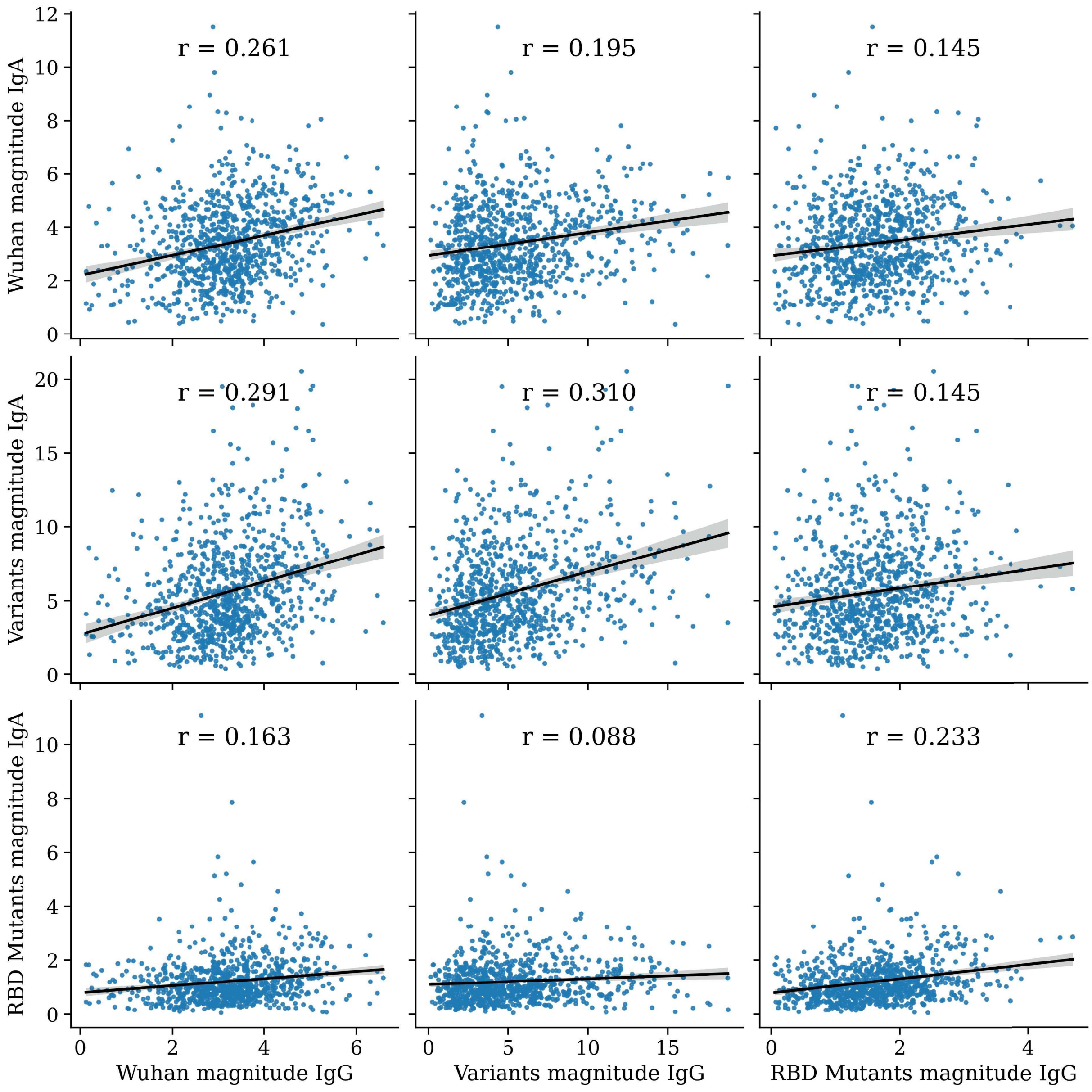
r = 0.261
r = 0.195
r = 0.145
r = 0.291
r = 0.310
r = 0.145
r = 0.163
r = 0.088
r = 0.233
Wuhan magnitude IgA
Variants magnitude IgA
RBD Mutants magnitude IgA
Wuhan magnitude IgG
Variants magnitude IgG
RBD Mutants magnitude IgG

**fig. S4: Correlation between IgG and IgA selected markers.** The correlation between the IgG and IgA markers that were used for combinations are presented. We used Pearson correlation coefficient to assess the association between the IgG and IgA markers.

**Table S1.**

| Description | virus | SARS-CoV-2 Variant | protein | magnitude columns | Source | Acknowledgment |
|---|---|---|---|---|---|---|
| CoV spike S1 SubunitHCoV-229E | 229E | - | S1 | - | Sino Biological, China | |
| CoV spike S1 Subunit, aa 1-760HCoV-HKU1 protein YP_173238.1 | HKU1 | - | S1 | - | Sino Biological, China | |
| CoV spike S1 Subunit HCoV-HKU1,isolate N5 (different strain) protein Q0ZME7.1 | HKU1 | - | S1 | - | Sino Biological, China | |
| CoV spike RBD, aa 367-606 MERS-CoV | MERS | - | RBD | - | Sino Biological, China | |
| CoV spike S1 Subunit, aa 1-725 MERS-CoV | MERS | - | S1 | - | Sino Biological, China | |
| CoV spike S1 SubunitHCoV-NL63 | NL63 | - | S1 | - | Sino Biological, China | |
| CoV spike S1+S2 HCoV-OC43 | OC43 | - | S1 and S2 | - | Sino Biological, China | |
| CoV spike RBDSARS-CoV, isolate WH20 | SARS-CoV | - | RBD | - | Sino Biological, China | |
| SARS-CoV spike S1 Subunit, isolate WH20 | SARS-CoV | - | S1 | - | Sino Biological, China | |

| | | | | | | |
|---|---|---|---|---|---|---|
| Spike Glycoprotein RBD from SARS-CoV-2, Alpha Variant with C Terminal Histidine Tag, Recombinant from HEK293 Cells | SARS-CoV-2 | Alpha lineage | RBD | - | BEI Resources, NIAID, NIH | The following reagent was obtained through BEI Resources, NIAID, NIH: Spike Glycoprotein Receptor Binding Domain (RBD) from SARS Related Coronavirus 2, Alpha Variant with C-Terminal Histidine Tag, Recombinant from HEK293 Cells, NR-55277. |
| Spike Glycoprotein RBD from SARS-CoV-2, Beta Variant with C Terminal Histidine Tag, Recombinant from HEK293 Cells | SARS-CoV-2 | Beta lineage | RBD | - | BEI Resources, NIAID, NIH | The following reagent was obtained through BEI Resources, NIAID, NIH: Spike Glycoprotein Receptor Binding Domain (RBD) from SARS Related Coronavirus 2, Beta Variant with C-Terminal Histidine Tag, Recombinant from HEK293 Cells, NR-55278. |

| | | | | | | |
|---|---|---|---|---|---|---|
| SARS-CoV-2 (2019-nCoV) Nucleocapsid Protein, YP_009724397.2(335 Gly/Ala) | SARS-CoV-2 | Wuhan lineage | NP | - | Sino Biological, China | |
| **Description** | **virus** | **SARS-CoV-2 Variant** | **protein** | **magnitude columns** | **Source** | **Acknowledgment** |
| Spike Glycoprotein RBD from SARS-CoV-2, L452R Variant with C-Terminal Histidine Tag, Recombinant from HEK293 Cells | SARS-CoV-2 | Point mutation_ NR-55403 | RBD | RBD_mutants | BEI Resources, NIAID, NIH | The following reagent was obtained through BEI Resources, NIAID, NIH: Spike Glycoprotein Receptor Binding Domain (RBD) from SARS-Related Coronavirus 2, L452R Variant with C-Terminal Histidine Tag, Recombinant from HEK293 Cells, NR-55403. |

| | | | | | | |
|---|---|---|---|---|---|---|
| Spike Glycoprotein RBD from SARS-CoV-2, N440K Variant with C-Terminal Histidine Tag, Recombinant from HEK293 Cells | SARS-CoV-2 | Point mutation_ NR-55405 | RBD | RBD_mutants | BEI Resources, NIAID, NIH | The following reagent was obtained through BEI Resources, NIAID, NIH: Spike Glycoprotein Receptor Binding Domain (RBD) from SARS-Related Coronavirus 2, N440K Variant with C-Terminal Histidine Tag, Recombinant from HEK293 Cells, NR-55405. |
| Spike Glycoprotein RBD from SARS-CoV-2, V483A Variant with C-Terminal Histidine Tag, Recombinant from HEK293 Cells | SARS-CoV-2 | Point mutation_ NR-55409 | RBD | RBD mutants | BEI Resources, NIAID, NIH | The following reagent was obtained through BEI Resources, NIAID, NIH: Spike Glycoprotein Receptor Binding Domain (RBD) from SARS-Related Coronavirus 2, V483A Variant with C-Terminal Histidine Tag, Recombinant from HEK293 Cells, NR-55409. |

| | | | | | | |
|---|---|---|---|---|---|---|
| Spike Glycoprotein RBD from SARS-CoV-2, K417N/E484K/N501Y Variant with C-Terminal Histidine Tag, Recombinant from HEK293 Cells | SARS-CoV-2 | Point mutation_ NR-55414 | RBD | RBD mutants | BEI Resources, NIAID, NIH | The following reagent was obtained through BEI Resources, NIAID, NIH: Spike Glycoprotein Receptor Binding Domain (RBD) from SARS-Related Coronavirus 2, K417N/E484K/ N501Y Variant with C-Terminal Histidine Tag, Recombinant from HEK293 Cells, NR-55414. |
| Spike Glycoprotein S1 Domain from SARS-CoV-2, Y144del Variant with C-Terminal Histidine Tag, Recombinant from HEK293 Cells | SARS-CoV-2 | Point mutation_ NR-55415 | S1 | S1 mutants | BEI Resources, NIAID, NIH | The following reagent was obtained through BEI Resources, NIAID, NIH: Spike Glycoprotein S1 Domain from SARS-Related Coronavirus 2, Y144del Variant with C-Terminal Histidine Tag, Recombinant from HEK293 Cells, NR-55415. |
| Spike Glycoprotein S1 Domain from SARS-CoV-2, HV69-70del Variant with C-Terminal Histidine Tag, Recombinant from HEK293 Cells | SARS-CoV-2 | Point mutation_ NR-55416 | S1 | S1_mutants | BEI Resources, NIAID, NIH | The following reagent was obtained through BEI Resources, NIAID, NIH: Spike Glycoprotein S1 Domain from SARS-Related |

| | | | | | | |
|---|---|---|---|---|---|---|
| | | | | | | Coronavirus 2, HV69-70del Variant with C-Terminal Histidine Tag, Recombinant from HEK293 Cells, NR-55416. |
| Spike Glycoprotein S1 Domain from SARS-CoV-2, D614G Variant with C-Terminal Histidine Tag, Recombinant from HEK293 Cells | SARS-CoV-2 | Point mutation_ NR-55418 | S1 | S1 mutants | BEI Resources, NIAID, NIH | The following reagent was obtained through BEI Resources, NIAID, NIH: Spike Glycoprotein S1 Domain from SARS-Related Coronavirus 2, D614G Variant with C-Terminal Histidine Tag, Recombinant from HEK293 Cells, NR-55418. |
| Spike Glycoprotein S1 Domain from SARS-CoV-2, P681H Variant with C-Terminal Histidine Tag, Recombinant from HEK293 Cells | SARS-CoV-2 | Point mutation_ NR-55420 | S1 | S1 mutants | BEI Resources, NIAID, NIH | The following reagent was obtained through BEI Resources, NIAID, NIH: Spike Glycoprotein S1 Domain from SARS-Related Coronavirus 2, P681H Variant with C-Terminal Histidine Tag, Recombinant from HEK293 Cells, NR-55420. |

| | | | | | | |
|---|---|---|---|---|---|---|
| Spike Glycoprotein (Stabilized) from SARS-CoV-2, B.1.1.1 Lineage with C-Terminal Histidine and Avi Tags, Recombinant from HEK293 Cells | SARS-CoV-2 | Alpha lineage | S1 and S2 | Variants | BEI Resources, NIAID, NIH | The following reagent was obtained through BEI Resources, NIAID, NIH: Spike Glycoprotein (Stabilized) from SARS-Related Coronavirus 2, B.1.1.1 Lineage with C-Terminal Histidine and Avi Tags, Recombinant from HEK293 Cells, NR-55615. |
| Spike Glycoprotein (Stabilized) from SARS-CoV-2, B.1.1.7 Lineage with C-Terminal Histidine and Avi Tags, Recombinant from HEK293 Cells | SARS-CoV-2 | Alpha lineage | S1 and S2 | Variants | BEI Resources, NIAID, NIH | The following reagent was obtained through BEI Resources, NIAID, NIH: Spike Glycoprotein (Stabilized) from SARS-Related Coronavirus 2, B.1.1.7 Lineage with C-Terminal Histidine and Avi Tags, Recombinant from HEK293 Cells, NR-55311. |

| | | | | | | |
|---|---|---|---|---|---|---|
| Spike Glycoprotein (Stabilized) from SARS-CoV-2, R.1 Lineage with C-Terminal Histidine and Avi Tags, Recombinant from HEK293 Cells | SARS-CoV-2 | Alpha lineage | S1 and S2 | Variants | BEI Resources, NIAID, NIH | The following reagent was obtained through BEI Resources, NIAID, NIH: Spike Glycoprotein (Stabilized) from SARS-Related Coronavirus 2, R.1 Lineage with C-Terminal Histidine and Avi Tags, Recombinant from HEK293 Cells, NR-55632. |
| Spike Glycoprotein (Stabilized) from SARS-CoV-2, B.1.351 Lineage with C-Terminal Histidine and Avi Tags, Recombinant from HEK293 Cells | SARS-CoV-2 | Beta lineage | S1 and S2 | Variants | BEI Resources, NIAID, NIH | The following reagent was obtained through BEI Resources, NIAID, NIH: Spike Glycoprotein (Stabilized) from SARS-Related Coronavirus 2, B.1.351 Lineage with C-Terminal Histidine and Avi Tags, Recombinant from HEK293 Cells, NR-55310. |

| | | | | | | |
|---|---|---|---|---|---|---|
| Spike Glycoprotein (Stabilized) from SARS-CoV-2, AY.2 Lineage (Delta Variant) with C-Terminal Histidine and Avi Tags, Recombinant from HEK293 Cells | SARS-CoV-2 | Delta and Kappa pre-lineage | S1 and S2 | Variants | BEI Resources, NIAID, NIH | The following reagent was obtained through BEI Resources, NIAID, NIH: Spike Glycoprotein (Stabilized) from SARS-Related Coronavirus 2, AY.2 Lineage (Delta Variant) with C-Terminal Histidine and Avi Tags, Recombinant from HEK293 Cells, NR-55711. |
| Spike Glycoprotein (Stabilized) from SARS-CoV-2, Delta Variant with C-Terminal Histidine and Avi Tags, Recombinant from HEK293 Cells | SARS-CoV-2 | Delta lineage | S1 and S2 | Variants | BEI Resources, NIAID, NIH | The following reagent was obtained through BEI Resources, NIAID, NIH: Spike Glycoprotein (Stabilized) from SARS-Related Coronavirus 2, Delta Variant with C-Terminal Histidine and Avi Tags, Recombinant from HEK293 Cells, NR-55614. |

| | | | | | | |
|---|---|---|---|---|---|---|
| Spike Glycoprotein (Stabilized) from SARS-CoV-2, A.23.1 Lineage with C-Terminal Histidine and Avi Tags, Recombinant from HEK293 Cells | SARS-CoV-2 | Delta sub-lineage | S1 and S2 | Variants | BEI Resources, NIAID, NIH | The following reagent was obtained through BEI Resources, NIAID, NIH: Spike Glycoprotein (Stabilized) from SARS-Related Coronavirus 2, A.23.1 Lineage with C-Terminal Histidine and Avi Tags, Recombinant from HEK293 Cells, NR-55616. |
| Spike Glycoprotein (Stabilized) from SARS-CoV-2, P.1 Lineage with C-Terminal Histidine and Avi Tags, Recombinant from HEK293 Cells | SARS-CoV-2 | Gamma lineage | S1 and S2 | Variants | BEI Resources, NIAID, NIH | The following reagent was obtained through BEI Resources, NIAID, NIH: Spike Glycoprotein (Stabilized) from SARS-Related Coronavirus 2, P.1 Lineage with C-Terminal Histidine and Avi Tags, Recombinant from HEK293 Cells, NR-55307. |

| | | | | | | |
|---|---|---|---|---|---|---|
| Spike Glycoprotein (Stabilized) from SARS-CoV-2, B.1.526 Lineage with C-Terminal Histidine and Avi Tags, Recombinant from HEK293 Cells | SARS-CoV-2 | Iota lineage | S1 and S2 | Variants | BEI Resources, NIAID, NIH | The following reagent was obtained through BEI Resources, NIAID, NIH: Spike Glycoprotein (Stabilized) from SARS-Related Coronavirus 2, B.1.526 Lineage with C-Terminal Histidine and Avi Tags, Recombinant from HEK293 Cells, NR-55438. |
| Spike Glycoprotein (Stabilized) from SARS-CoV-2, Kappa Variant with C-Terminal Histidine and Avi Tags, Recombinant from HEK293 Cells | SARS-CoV-2 | Kappa lineage | S1 and S2 | Variants | BEI Resources, NIAID, NIH | The following reagent was obtained through BEI Resources, NIAID, NIH: Spike Glycoprotein (Stabilized) from SARS-Related Coronavirus 2, Kappa Variant with C-Terminal Histidine and Avi Tags, Recombinant from HEK293 Cells, NR-55495. |

| | | | | | | |
|---|---|---|---|---|---|---|
| Spike Glycoprotein (Stabilized) from SARS-CoV-2, B.1.621 Lineage (Mu Variant) with C-Terminal Histidine and Avi Tags, Recombinant from HEK293 Cells | SARS-CoV-2 | Mu lineage | S1 and S2 | Variants | BEI Resources, NIAID, NIH | The following reagent was obtained through BEI Resources, NIAID, NIH: Spike Glycoprotein (Stabilized) from SARS-Related Coronavirus 2, B.1.621 Lineage (Mu Variant) with C-Terminal Histidine and Avi Tags, Recombinant from HEK293 Cells, NR-55712. |
| Spike Glycoprotein (Stabilized) from SARS-CoV-2, Theta Variant with C-Terminal Histidine and Avi Tags, Recombinant from HEK293 Cells | SARS-CoV-2 | Theta linage | S1 and S2 | Variants | BEI Resources, NIAID, NIH | The following reagent was obtained through BEI Resources, NIAID, NIH: Spike Glycoprotein (Stabilized) from SARS-Related Coronavirus 2, Theta Variant with C-Terminal Histidine and Avi Tags, Recombinant from HEK293 Cells, NR-55633. |

| | | | | | | |
|---|---|---|---|---|---|---|
| Spike Glycoprotein RBD from SARS-CoV-2, Wuhan-Hu-1 with C-Terminal Histidine Tag, Recombinant from HEK293 Cells (This item replaces NR-52306) | SARS-CoV-2 | Wuhan lineage | RBD | Wuhan | BEI Resources, NIAID, NIH | The following reagent was produced under HHSN272201400008C and obtained through BEI Resources, NIAID, NIH: Spike Glycoprotein Receptor Binding Domain (RBD) from SARS-Related Coronavirus 2, Wuhan-Hu-1 with C-Terminal Histidine Tag, Recombinant from HEK293F Cells, NR-52366. |
| Spike Glycoprotein (Stabilized) from SARS-CoV-2, Wuhan-Hu-1 with C-Terminal Histidine Tag, Recombinant from HEK293F Cells | SARS-CoV-2 | Wuhan lineage | S1 and S2 | Wuhan | BEI Resources, NIAID, NIH | The following reagent was produced under HHSN272201400008C and obtained through BEI Resources, NIAID, NIH: Spike Glycoprotein (Stabilized) from SARS-Related Coronavirus 2, Wuhan-Hu-1 with C-Terminal Histidine Tag, Recombinant from HEK293F Cells, NR-52397. |

| | | | | | |
|---|---|---|---|---|---|
| SARS-CoV-2 (2019-nCoV) Spike S1, | SAR S-CoV -2 | Wuhan lineage | S1 | Wuhan | Sino Biologi cal, China |
| produced in human HEK293 cells<br>SARS-CoV-2 (2019-nCoV) Spike RBD, | SAR S-CoV -2<br>SAR S-CoV -2 | Wuhan lineage<br>Wuhan lineage | S1<br>RBD | Wuhan<br>- | Sino Biologi cal, China<br>Sino Biologi cal, China |
| produced in baculovirus-infected insect cells<br>SARS-CoV-2 (2019-nCoV) Spike RBD, | SAR S-CoV -2<br>SAR S-CoV -2 | Wuhan lineage<br>Wuhan lineage | RBD<br>RBD | -<br>Wuhan | Sino Biologi cal, China<br>Sino Biologi cal, China |
| produced in human HEK293 cells | SAR S-CoV -2 | Wuhan lineage | RBD | Wuhan | Sino Biologi cal, China |

**Table S2.**

Characteristics of the 74 individuals in the immunogenicity subset used in the study, which included 38 low-baseline and 36 high-baseline individuals**.**

| | **Overall** | **high** | **low** | **p-value** |
|---|---|---|---|---|
| N | 74 | 36 | 38 | |
| Second booster (%) | 58 (78.4) | 29 (80.6) | 29 (76.3) | 0.873 |
| Positive PCR 60-90 follow up days (%) | 31 (41.9) | 11 (30.6) | 20 (52.6) | 0.091 |
| Positive PCR 30 follow up days (%) | 23 (31.1) | 7 (19.4) | 16 (42.1) | 0.064 |
| Sex, Male (%) | 27 (36.5) | 18 (50.0) | 9 (23.7) | 0.035 |
| Age (mean (SD)) | 46.56 (12.35) | 47.34 (13.45) | 45.82 (11.33) | 0.6 |
| Age group(%) | | | | 0.822 |
| 18-34 | 14 (18.9) | 7 (19.4) | 7 (18.4) | |
| 35-49 | 30 (40.5) | 14 (38.9) | 16 (42.1) | |
| 50-64 | 24 (32.4) | 11 (30.6) | 13 (34.2) | |
| 65+ | 6 (8.1) | 4 (11.1) | 2 (5.3) | |
| Ethnicity (%) | | | | NaN |
| Jewish | 71 (98.6) | 33 (97.1) | 38 (100.0) | |
| Bedouin | 1 (1.4) | 1 (2.9) | 0 (0.0) | |
| Other or Not Mentioned | 0 (0.0) | 0 (0.0) | 0 (0.0) | |
| Socioeconomic status (%) | | | | NaN |
| Very High | 15 (20.3) | 8 (22.2) | 7 (18.4) | |
| High | 27 (36.5) | 15 (41.7) | 12 (31.6) | |
| Medium | 24 (32.4) | 10 (27.8) | 14 (36.8) | |
| Low | 5 (6.8) | 2 (5.6) | 3 (7.9) | |
| Very Low | 0 (0.0) | 0 (0.0) | 0 (0.0) | |
| no data | 3 (4.1) | 1 (2.8) | 2 (5.3) | |
| Occuppation (%) | | | | 0.325 |
| Physician | 21 (28.4) | 13 (36.1) | 8 (21.1) | |

| | | | | |
|---|---|---|---|---|
| Nurse | 19 (25.7) | 9 (25.0) | 10 (26.3) | |
| Administration and support staff | 34 (45.9) | 14 (38.9) | 20 (52.6) | |
| Medical Center, Soroka (%) | 74 (100.0) | 36 (100.0) | 38 (100.0) | NA |
| Days since third dose (mean (SD)) | 143.01 (6.62) | 143.75 (6.09) | 142.32 (7.09) | 0.355 |
| PCR test count (median [IQR]) | 1.00 [0.00, 3.00] | 1.00 [0.00, 3.00] | 2.00 [0.25, 3.00] | 0.5 |
| Daily interaction with corona patients (%) | | | | 0.093 |
| Yes | 12 (16.2) | 9 (25.0) | 3 (7.9) | |
| No | 61 (82.4) | 27 (75.0) | 34 (89.5) | |
| Not Mentioned | 1 (1.4) | 0 (0.0) | 1 (2.6) | |

**Table S3.**

Results of Cox proportional hazard model estimating vaccine efficacy for 30 follow up days. The model used calendar days as the time-axis and was adjusted for age, occupation, medical center, and time from the third vaccination.

| Variable | Hazard Ratio | lower .95 | upper .95 | P-value |
|---|---|---|---|---|
| vaccinated Yes | 0.55 | 0.37 | 0.81 | 0.002 |
| gender Male | 0.98 | 0.67 | 1.43 | 0.91 |
| age grp 35-49 | 1.15 | 0.71 | 1.85 | 0.576 |
| age grp 50-64 | 0.8 | 0.48 | 1.32 | 0.382 |
| age grp 65+ | 0.51 | 0.19 | 1.35 | 0.174 |
| sector of occuppation grp Physicians or Nurses | 1.03 | 0.75 | 1.43 | 0.842 |
| hospital Emek | 1.16 | 0.65 | 2.06 | 0.61 |
| hospital Meir | 0.89 | 0.42 | 1.86 | 0.749 |
| hospital Soroka | 1.03 | 0.61 | 1.71 | 0.923 |
| time from third vaccine (months) | 1.03 | 0.83 | 1.27 | 0.792 |

**Table S4.**

Results of Cox proportional hazard model estimating vaccine efficacy at 60-90 follow up days. The model used calendar days as the time-axis and was adjusted for age, occupation, medical center, and time from the third vaccination.

| Variable | Hazard Ratio | lower .95 | upper .95 | P-value |
|---|---|---|---|---|
| vaccinated Yes | 0.63 | 0.46 | 0.85 | 0.003 |
| gender Male | 0.88 | 0.65 | 1.21 | 0.439 |
| age grp 35-49 | 1.06 | 0.72 | 1.56 | 0.755 |
| age grp 50-64 | 0.76 | 0.51 | 1.13 | 0.174 |
| age grp 65+ | 0.44 | 0.2 | 0.97 | 0.041 |
| sector of occuppation grp Physicians or Nurses | 1 | 0.77 | 1.3 | 0.989 |
| hospital Emek | 1.1 | 0.69 | 1.76 | 0.696 |
| hospital Meir | 1.16 | 0.66 | 2.04 | 0.597 |
| hospital Soroka | 1.06 | 0.7 | 1.59 | 0.792 |
| time from third vaccine (months) | 1.06 | 0.89 | 1.27 | 0.525 |

**Table S5.**

Results of Poisson regression estimating vaccine efficacy for 30 follow up days. The model was adjusted to the daily proportion of positive PCR tests to Covid-19, age, occupation, medical center, and time from the third vaccination and subjects as a random effect.

| **Variable** | **Incidence rate ratio** | **lower .95** | **upper .95** | **p-value** |
|---|---|---|---|---|
| vaccinated Yes | 0.43 | 0.28 | 0.64 | <0.001 |
| percent pcr | 1.14 | 1.09 | 1.18 | <0.001 |
| age grp 35-49 | 1.12 | 0.68 | 1.82 | 0.66 |
| age grp 50-64 | 0.8 | 0.47 | 1.34 | 0.387 |
| age grp 65+ | 0.45 | 0.15 | 1.33 | 0.151 |
| hospital Emek | 1.28 | 0.69 | 2.35 | 0.431 |
| hospital Meir | 1.01 | 0.47 | 2.19 | 0.972 |
| hospital Soroka | 1.23 | 0.7 | 2.13 | 0.471 |
| sector of occuppation grp Physicians or Nurses | 1.13 | 0.81 | 1.59 | 0.467 |
| time from third vaccine (months) | 1.02 | 0.82 | 1.25 | 0.886 |
| gender Male | 0.99 | 0.67 | 1.47 | 0.96 |

**Table S6.**

Results of Poisson regression estimating vaccine efficacy for 60-90 follow up days. The model was adjusted to the daily proportion of positive PCR tests to Covid-19, age, occupation, medical center, and time from the third vaccination and subjects as a random effect.

| Variable | Incidence rate ratio | lower .95 | upper .95 | p-value |
|---|---|---|---|---|
| vaccinated Yes | 0.55 | 0.41 | 0.74 | <0.001 |
| percent pcr | 1.08 | 1.06 | 1.1 | <0.001 |
| age grp 35-49 | 1.08 | 0.74 | 1.59 | 0.683 |
| age grp 50-64 | 0.77 | 0.52 | 1.16 | 0.21 |
| age grp 65+ | 0.47 | 0.21 | 1.01 | 0.054 |
| hospital Emek | 1.06 | 0.66 | 1.7 | 0.804 |
| hospital Meir | 1.12 | 0.64 | 1.96 | 0.686 |
| hospital Soroka | 1.08 | 0.72 | 1.62 | 0.714 |
| sector of occuppation grp Physicians or Nurses | 1.02 | 0.79 | 1.33 | 0.874 |
| time from third vaccine (months) | 1.06 | 0.89 | 1.27 | 0.509 |
| gender Male | 0.88 | 0.64 | 1.2 | 0.406 |

**Table S7.**

Results of Cox proportional hazard model comparing infection hazard at 30 follow up days in the low-baseline with high-baseline response groups, using the five primary analysis baseline markers. The model used calendar days as the time-axis and was adjusted for age, occupation, medical center, and time from the third vaccination.

| Marker | Hazard Ratio | lower .95 | upper .95 | p-value |
|---|---|---|---|---|
| **Three doses** | | | | |
| IgG BioPlex S2 | 1.39 | 0.95 | 2.05 | 0.089 |
| IgG Alinity RBD | 1.59 | 1.07 | 2.35 | 0.02 |
| IgG Mutants RBD | 1.5 | 0.87 | 2.61 | 0.148 |
| IgA Variants | 1.34 | 0.76 | 2.37 | 0.309 |
| IgA Wuhan | 1.09 | 0.63 | 1.89 | 0.756 |
| **Four doses** | | | | |
| IgG BioPlex S2 | 1.32 | 0.71 | 2.45 | 0.389 |
| IgG Alinity RBD | 1.47 | 0.77 | 2.78 | 0.239 |
| IgG Mutants RBD | 2.07 | 0.73 | 5.83 | 0.171 |
| IgA Variants | 4.45 | 1.52 | 13.02 | 0.006 |
| IgA Wuhan | 3.19 | 1.21 | 8.38 | 0.019 |

**Table S8.**

Results of Cox proportional hazard model comparingInfection hazard at 60-90 follow up days In the low-baseline with high-baseline response groups, using the five primary analysis baseline markers. The model used calendar days as the time-axis and was adjusted for age, occupation, medical center, and time from the third vaccination.

| **Marker** | **Hazard Ratio** | **lower .95** | **upper .95** | **p-value** |
|---|---|---|---|---|
| **Three doses** | | | | |
| IgG BioPlex S2 | 1.45 | 1.05 | 1.99 | 0.022 |
| IgG Alinity RBD | 1.54 | 1.12 | 2.13 | 0.008 |
| IgG Mutants RBD | 1.52 | 0.95 | 2.42 | 0.079 |
| IgA Variants | 1.47 | 0.92 | 2.34 | 0.107 |
| IgA Wuhan | 1.33 | 0.84 | 2.11 | 0.217 |
| **Four doses** | | | | |
| IgG BioPlex S2 | 1.77 | 1.07 | 2.92 | 0.025 |
| IgG Alinity RBD | 1.65 | 1 | 2.71 | 0.049 |
| IgG Mutants RBD | 2.24 | 0.99 | 5.04 | 0.053 |
| IgA Variants | 2.04 | 0.96 | 4.35 | 0.065 |
| IgA Wuhan | 2.05 | 1.03 | 4.09 | 0.041 |

**Table S9.**

Results of Cox proportional hazard model comparing infection hazard at 60-90 follow up days in the low-baseline with high-baseline response groups using all markers. The model estimated HR for the individuals receiving three doses of the Pfizer vaccine. We used calendar days as the time-axis and adjusted for age, occupation, medical center, and time from the third vaccination. Results presented in a decreasing order based on HR value, significant p-values are colored in green.

| Marker | Hazard Ratio | lower .95 | upper .95 | p-value |
|---|---|---|---|---|
| SARS CoV 2 S R.1 IgG | 2.14 | 1.19 | 3.85 | **0.011** |
| SARS CoV 2 RBD IgA | 2.03 | 1.29 | 3.2 | **0.002** |
| SARS CoV 2 S1 IgA | 2 | 1.19 | 3.36 | **0.009** |
| SARS CoV 2 RBD N440K IgA | 1.92 | 1.22 | 3.04 | **0.005** |
| SARS CoV 2 S B.1.1.7 IgG | 1.88 | 1.05 | 3.37 | **0.034** |
| SARS1 RBD IgA | 1.83 | 1.16 | 2.9 | **0.009** |
| SARS CoV 2 S B.1.351 IgG | 1.79 | 1.03 | 3.1 | **0.039** |
| SARS S1 IgA | 1.72 | 1.08 | 2.73 | **0.022** |
| SARS CoV 2 RBD Wuhan Hu 1 IgA | 1.68 | 1.07 | 2.65 | **0.025** |
| SARS CoV 2 S B.1.1.7 IgA | 1.68 | 1.06 | 2.66 | **0.029** |
| SARS CoV 2 S1 D614G IgG | 1.63 | 0.93 | 2.87 | 0.088 |
| SARS CoV 2 S1 Y144del IgG | 1.62 | 0.95 | 2.75 | 0.076 |
| SARS CoV 2 S B.1.351 IgA | 1.62 | 1.03 | 2.55 | **0.038** |
| SARS CoV 2 RBD Wuhan Hu 1 IgG | 1.6 | 0.9 | 2.85 | 0.11 |
| SARS CoV 2 S AY.2 IgA | 1.6 | 1 | 2.56 | 0.051 |
| SARS CoV 2 S R.1 IgA | 1.58 | 0.99 | 2.53 | 0.054 |
| SARS CoV 2 S Theta IgA | 1.58 | 1 | 2.48 | **0.049** |
| SARS CoV 2 S B.1.621 IgA | 1.56 | 1 | 2.44 | 0.052 |
| SARS CoV 2 S B.1.526 IgA | 1.5 | 0.96 | 2.34 | 0.076 |
| SARS CoV 2 S P1 IgA | 1.49 | 0.92 | 2.41 | 0.107 |
| SARS CoV 2 S Delta IgG | 1.47 | 0.86 | 2.53 | 0.158 |
| SARS CoV 2 S B.1.1.1 IgA | 1.47 | 0.92 | 2.35 | 0.107 |
| SARS CoV 2 S B.1.526 IgG | 1.43 | 0.81 | 2.51 | 0.218 |
| SARS CoV 2 S B.1.1.1 IgG | 1.42 | 0.79 | 2.55 | 0.239 |
| SARS CoV 2 RBD V483A IgG | 1.42 | 0.81 | 2.47 | 0.219 |
| SARS CoV 2 S P1 IgG | 1.39 | 0.76 | 2.54 | 0.279 |
| SARS CoV 2 S1 Y144del IgA | 1.38 | 0.8 | 2.4 | 0.25 |
| SARS CoV 2 S Kappa IgA | 1.36 | 0.87 | 2.14 | 0.175 |
| SARS CoV 2 RBD Alpha IgA | 1.33 | 0.85 | 2.06 | 0.211 |
| MERS S1 IgG | 1.3 | 0.82 | 2.04 | 0.26 |
| SARS CoV 2 RBD Alpha IgG | 1.28 | 0.79 | 2.08 | 0.312 |
| SARS CoV 2 RBD IgG | 1.28 | 0.82 | 2.01 | 0.282 |
| SARS CoV 2 S Theta IgG | 1.28 | 0.66 | 2.47 | 0.467 |

| | | | | |
|---|---|---|---|---|
| SARS CoV 2 S1 D614G IgA | 1.26 | 0.76 | 2.09 | 0.379 |
| SARS CoV 2 RBD L452R IgA | 1.24 | 0.8 | 1.92 | 0.346 |
| SARS CoV 2 S1 HV69 70del IgA | 1.23 | 0.78 | 1.95 | 0.368 |
| SARS CoV 2 S Delta IgA | 1.21 | 0.78 | 1.89 | 0.396 |
| SARS CoV 2 S1 IgG | 1.21 | 0.76 | 1.91 | 0.422 |
| SARS1 RBD IgG | 1.2 | 0.73 | 1.99 | 0.465 |
| SARS CoV 2 RBD N440K IgG | 1.19 | 0.77 | 1.83 | 0.434 |
| SARS CoV 2 RBD V483A IgA | 1.17 | 0.68 | 1.99 | 0.572 |
| SARS CoV 2 S B.1.621 IgG | 1.17 | 0.64 | 2.14 | 0.62 |
| SARS CoV 2 S Kappa IgG | 1.15 | 0.64 | 2.06 | 0.644 |
| SARS CoV 2 S A.23.1 IgG | 1.14 | 0.55 | 2.34 | 0.726 |
| MERS S1 IgA | 1.13 | 0.73 | 1.74 | 0.576 |
| SARS CoV 2 RBD K417N/E484K/N501Y IgG | 1.12 | 0.57 | 2.19 | 0.742 |
| SARS CoV 2 RBD K417N/E484K/N501Y IgA | 1.11 | 0.71 | 1.73 | 0.645 |
| SARS CoV 2 RBD L452R IgG | 1.1 | 0.68 | 1.78 | 0.69 |
| SARS CoV 2 S Wuhan Hu 1 IgG | 1.08 | 0.67 | 1.74 | 0.748 |
| MERS RBD IgG | 1.07 | 0.68 | 1.7 | 0.765 |
| SARS CoV 2 RBD Beta IgA | 1.06 | 0.66 | 1.7 | 0.819 |
| SARS CoV 2 S Wuhan Hu 1 IgA | 1.04 | 0.65 | 1.65 | 0.878 |
| SARS S1 IgG | 1.03 | 0.65 | 1.63 | 0.91 |
| SARS CoV 2 S1 P681H IgG | 1 | 0.59 | 1.69 | 0.997 |
| SARS CoV 2 S A.23.1 IgA | 0.98 | 0.58 | 1.66 | 0.943 |
| SARS CoV 2 S AY.2 IgG | 0.97 | 0.53 | 1.8 | 0.932 |
| RSV G IgA | 0.97 | 0.6 | 1.55 | 0.885 |
| NL63 S1 IgG | 0.95 | 0.59 | 1.52 | 0.83 |
| SARS CoV 2 S1 P681H IgA | 0.94 | 0.59 | 1.51 | 0.811 |
| SARS CoV 2 NP IgG | 0.92 | 0.49 | 1.75 | 0.803 |
| SARS CoV 2 NP IgA | 0.9 | 0.57 | 1.43 | 0.668 |
| SARS CoV 2 S1 HV69 70del IgG | 0.89 | 0.57 | 1.41 | 0.621 |
| OC43 S IgG | 0.85 | 0.54 | 1.32 | 0.468 |
| HKU1 S1 IgA | 0.83 | 0.54 | 1.28 | 0.41 |
| 229E S1 IgG | 0.8 | 0.5 | 1.28 | 0.351 |

| | | | | |
|---|---|---|---|---|
| HKU1 S1 IgG | 0.74 | 0.47 | 1.18 | 0.212 |
| NL63 S1 IgA | 0.73 | 0.46 | 1.16 | 0.187 |
| OC43 S IgA | 0.7 | 0.44 | 1.11 | 0.131 |
| SARS CoV 2 RBD Beta IgG | 0.65 | 0.36 | 1.17 | 0.15 |
| RSV G IgG | 0.64 | 0.32 | 1.28 | 0.204 |
| 229E S1 IgA | 0.63 | 0.4 | 1 | 0.051 |

**Table S10.**

Results of Cox proportional hazard model comparing infection hazard at 60-90 follow up days in the low-baseline with high-baseline response groups using all markers. The model estimated HR for the individuals receiving four doses of the Pfizer vaccine. We used calendar days as the time-axis and adjusted for age, occupation, medical center, and time from the third vaccination. Results presented in a decreasing order based on HR value, significant p-values are colored in green.

| Marker | Hazard Ratio | lower .95 | upper .95 | p-value |
|---|---|---|---|---|
| SARS CoV 2 S B.1.621 IgA | 2.82 | 1.22 | 6.54 | **0.015** |
| SARS CoV 2 S Delta IgA | 2.3 | 1.11 | 4.76 | **0.025** |
| SARS CoV 2 S1 P681H IgG | 2.27 | 1.01 | 5.13 | **0.048** |
| SARS CoV 2 RBD K417N/E484K/N501Y IgG | 2.27 | 0.89 | 5.76 | 0.085 |
| SARS CoV 2 S P1 IgA | 2.2 | 0.97 | 4.99 | 0.059 |
| SARS CoV 2 RBD N440K IgA | 2.18 | 1.05 | 4.53 | **0.036** |
| SARS CoV 2 RBD V483A IgG | 2.16 | 1.04 | 4.48 | **0.04** |
| SARS CoV 2 RBD N440K IgG | 2.12 | 0.93 | 4.83 | 0.075 |
| SARS CoV 2 RBD Wuhan Hu 1 IgA | 2.11 | 0.94 | 4.72 | 0.07 |
| SARS CoV 2 RBD IgG | 2.1 | 1 | 4.4 | **0.049** |
| SARS CoV 2 RBD L452R IgA | 2.03 | 0.97 | 4.26 | 0.061 |
| SARS CoV 2 S1 HV69 70del IgG | 2.02 | 0.97 | 4.22 | 0.06 |
| SARS CoV 2 S Kappa IgA | 1.96 | 0.94 | 4.06 | 0.071 |
| SARS CoV 2 RBD IgA | 1.9 | 0.91 | 3.96 | 0.088 |
| SARS CoV 2 S B.1.1.1 IgG | 1.9 | 0.81 | 4.46 | 0.143 |
| SARS CoV 2 S Kappa IgG | 1.88 | 0.81 | 4.35 | 0.14 |
| SARS CoV 2 RBD Wuhan Hu 1 IgG | 1.88 | 0.84 | 4.23 | 0.126 |
| SARS CoV 2 S1 IgG | 1.84 | 0.94 | 3.61 | 0.076 |
| SARS CoV 2 S A.23.1 IgA | 1.77 | 0.81 | 3.85 | 0.149 |
| SARS CoV 2 RBD Alpha IgG | 1.67 | 0.82 | 3.39 | 0.154 |
| SARS CoV 2 S AY.2 IgA | 1.65 | 0.81 | 3.37 | 0.167 |
| SARS CoV 2 NP IgG | 1.64 | 0.66 | 4.07 | 0.289 |
| SARS CoV 2 S R.1 IgG | 1.62 | 0.7 | 3.76 | 0.26 |
| SARS S1 IgA | 1.61 | 0.74 | 3.52 | 0.231 |
| SARS CoV 2 RBD L452R IgG | 1.58 | 0.82 | 3.05 | 0.17 |
| SARS CoV 2 S Theta IgA | 1.58 | 0.76 | 3.25 | 0.217 |
| SARS CoV 2 S B.1.526 IgG | 1.56 | 0.62 | 3.91 | 0.343 |
| SARS CoV 2 S1 D614G IgG | 1.53 | 0.63 | 3.7 | 0.347 |
| SARS CoV 2 RBD Alpha IgA | 1.51 | 0.71 | 3.21 | 0.28 |
| SARS CoV 2 S A.23.1 IgG | 1.48 | 0.61 | 3.58 | 0.388 |
| SARS CoV 2 S1 D614G IgA | 1.47 | 0.67 | 3.24 | 0.338 |
| SARS CoV 2 S B.1.526 IgA | 1.45 | 0.7 | 2.99 | 0.312 |

| | | | | |
|---|---|---|---|---|
| SARS CoV 2 S1 Y144del IgG | 1.44 | 0.64 | 3.27 | 0.379 |
| SARS CoV 2 S1 HV69 70del IgA | 1.43 | 0.69 | 2.97 | 0.335 |
| SARS CoV 2 S AY.2 IgG | 1.43 | 0.63 | 3.24 | 0.393 |
| MERS RBD IgA | 1.38 | 0.71 | 2.68 | 0.339 |
| SARS CoV 2 S B.1.1.1 IgA | 1.37 | 0.67 | 2.84 | 0.391 |
| SARS CoV 2 S B.1.351 IgA | 1.35 | 0.63 | 2.88 | 0.435 |
| MERS S1 IgG | 1.34 | 0.66 | 2.74 | 0.416 |
| SARS CoV 2 S Wuhan Hu 1 IgA | 1.27 | 0.6 | 2.69 | 0.529 |
| SARS CoV 2 S B.1.1.7 IgG | 1.27 | 0.52 | 3.07 | 0.6 |
| SARS CoV 2 S B.1.1.7 IgA | 1.26 | 0.58 | 2.73 | 0.561 |
| SARS CoV 2 S P1 IgG | 1.24 | 0.5 | 3.06 | 0.645 |
| SARS CoV 2 S Delta IgG | 1.22 | 0.62 | 2.38 | 0.568 |
| SARS CoV 2 S1 IgA | 1.21 | 0.6 | 2.47 | 0.596 |
| SARS CoV 2 RBD Beta IgA | 1.2 | 0.57 | 2.53 | 0.627 |
| SARS CoV 2 RBD K417N/E484K/N501Y IgA | 1.17 | 0.58 | 2.34 | 0.659 |
| SARS CoV 2 S B.1.351 IgG | 1.14 | 0.46 | 2.81 | 0.774 |
| SARS CoV 2 S1 Y144del IgA | 1.11 | 0.47 | 2.62 | 0.807 |
| SARS1 RBD IgA | 1.08 | 0.52 | 2.24 | 0.829 |
| SARS CoV 2 RBD V483A IgA | 1.04 | 0.5 | 2.15 | 0.916 |
| NL63 S1 IgA | 1.02 | 0.52 | 1.99 | 0.955 |
| SARS CoV 2 RBD Beta IgG | 1.02 | 0.47 | 2.22 | 0.963 |
| SARS CoV 2 S B.1.621 IgG | 1.01 | 0.43 | 2.38 | 0.98 |
| 229E S1 IgA | 0.97 | 0.51 | 1.83 | 0.916 |
| SARS CoV 2 S Theta IgG | 0.94 | 0.35 | 2.51 | 0.9 |
| MERS RBD IgG | 0.94 | 0.48 | 1.84 | 0.848 |
| OC43 S IgA | 0.86 | 0.45 | 1.67 | 0.663 |
| NL63 S1 IgG | 0.84 | 0.41 | 1.74 | 0.645 |
| SARS CoV 2 S R.1 IgA | 0.84 | 0.41 | 1.74 | 0.641 |
| MERS S1 IgA | 0.75 | 0.36 | 1.53 | 0.427 |
| 229E S1 IgG | 0.73 | 0.32 | 1.65 | 0.447 |
| RSV G IgA | 0.72 | 0.36 | 1.45 | 0.355 |
| SARS CoV 2 S Wuhan Hu 1 IgG | 0.68 | 0.31 | 1.5 | 0.34 |
| OC43 S IgG | 0.66 | 0.34 | 1.29 | 0.227 |
| SARS CoV 2 S1 P681H IgA | 0.63 | 0.26 | 1.52 | 0.304 |

| | | | | |
|---|---|---|---|---|
| HKU1 S1 IgG | 0.57 | 0.25 | 1.26 | 0.163 |
| SARS S1 IgG | 0.53 | 0.25 | 1.11 | 0.091 |
| SARS1 RBD IgG | 0.53 | 0.22 | 1.24 | 0.143 |
| HKU1 S1 IgA | 0.4 | 0.19 | 0.81 | **0.011** |
| RSV G IgG | 0.35 | 0.14 | 0.87 | **0.025** |

**Table S11.**

Results of Cox proportional hazard model comparing infection hazard at 30 follow up days in the low-baseline with high-baseline response groups, using all pairwise combinations of the five primary analysis baseline markers. The model used calendar days as the time-axis and was adjusted for age, occupation, medical center, and time from the third vaccination.

| **Marker combination** | **Hazard Ratio** | **lower .95** | **upper .95** | **p-value** |
|---|---|---|---|---|
| **Three doses** | | | | |
| IgG Mutants RBD & IgA Variants | 4.49 | 1.07 | 18.8 | 0.04 |
| IgG Alinity RBD & IgA Variants | 3.45 | 1.35 | 8.81 | 0.01 |
| IgG Alinity RBD & IgG Mutants RBD | 1.98 | 1.05 | 3.7 | 0.034 |
| IgG Mutants RBD & IgA Wuhan | 2.75 | 0.74 | 10.2 | 0.13 |
| BioPlex IgG S2 & IgG Alinity RBD | 1.76 | 1.11 | 2.79 | 0.016 |
| BioPlex IgG S2 & IgG Mutants RBD | 1.93 | 0.99 | 3.77 | 0.055 |
| BioPlex IgG S2 & IgA Variants | 2.94 | 1.22 | 7.07 | 0.016 |
| BioPlex IgG S2 & IgA Wuhan | 1.63 | 0.72 | 3.67 | 0.237 |
| IgA Wuhan & IgA Variants | 1.48 | 0.73 | 3.02 | 0.277 |
| IgG Alinity RBD & IgA Wuhan | 1.64 | 0.72 | 3.76 | 0.242 |
| **Four doses** | | | | |
| IgG Mutants RBD & IgA Variants | 12.24 | 1.15 | 130.11 | 0.038 |
| IgG Alinity RBD & IgA Variants | 4.38 | 1.23 | 15.65 | 0.023 |
| IgG Alinity RBD & IgG Mutants RBD | 1.96 | 0.6 | 6.42 | 0.269 |
| IgG Mutants RBD & IgA Wuhan | 6.15 | 0.94 | 40.38 | 0.058 |
| BioPlex IgG S2 & IgG Alinity RBD | 1.72 | 0.76 | 3.9 | 0.195 |
| BioPlex IgG S2 & IgG Mutants RBD | 2.77 | 0.69 | 11.16 | 0.153 |
| BioPlex IgG S2 & IgA Variants | 3.04 | 0.92 | 10.07 | 0.068 |
| BioPlex IgG S2 & IgA Wuhan | 2.87 | 0.88 | 9.38 | 0.082 |
| IgA Wuhan & IgA Variants | 5.73 | 1.54 | 21.26 | 0.009 |
| IgG Alinity RBD & IgA Wuhan | 2.51 | 0.82 | 7.74 | 0.108 |

**Table S12.**

Results of Cox proportional hazard model comparing infection hazard at 60-90 follow up days in the low-baseline with high-baseline response groups, using all pairwise combinations of the five primary analysis baseline markers. The model used calendar days as the time-axis and was adjusted for age, occupation, medical center, and time from the third vaccination.

| **Marker combination** | **Hazard Ratio** | **lower .95** | **upper .95** | **p-value** |
|---|---|---|---|---|
| **Three doses** | | | | |
| IgG Mutants RBD & IgA Variants | 6.34 | 1.62 | 24.86 | 0.008 |
| IgG Alinity RBD & IgA Variants | 3.3 | 1.5 | 7.25 | 0.003 |
| IgG Alinity RBD & IgG Mutants RBD | 1.9 | 1.12 | 3.22 | 0.017 |
| IgG Mutants RBD & IgA Wuhan | 4.94 | 1.39 | 17.58 | 0.014 |
| BioPlex IgG S2 & IgG Alinity RBD | 1.84 | 1.24 | 2.72 | 0.002 |
| BioPlex IgG S2 & IgG Mutants RBD | 1.76 | 1.01 | 3.07 | 0.046 |
| BioPlex IgG S2 & IgA Variants | 2.88 | 1.32 | 6.28 | 0.008 |
| BioPlex IgG S2 & IgA Wuhan | 2.23 | 1.08 | 4.59 | 0.03 |
| IgA Wuhan & IgA Variants | 1.58 | 0.88 | 2.82 | 0.123 |
| IgG Alinity RBD & IgA Wuhan | 1.95 | 0.97 | 3.95 | 0.062 |
| **Four doses** | | | | |
| IgG Mutants RBD & IgA Variants | 8.14 | 1.43 | 46.41 | 0.018 |
| IgG Alinity RBD & IgA Variants | 3.47 | 1.23 | 9.84 | 0.019 |
| IgG Alinity RBD & IgG Mutants RBD | 2.27 | 0.89 | 5.83 | 0.088 |
| IgG Mutants RBD & IgA Wuhan | 7.67 | 1.62 | 36.29 | 0.01 |
| BioPlex IgG S2 & IgG Alinity RBD | 2.01 | 1.09 | 3.7 | 0.025 |
| BioPlex IgG S2 & IgG Mutants RBD | 2.7 | 0.94 | 7.75 | 0.066 |
| BioPlex IgG S2 & IgA Variants | 2.61 | 0.95 | 7.17 | 0.064 |
| BioPlex IgG S2 & IgA Wuhan | 3.29 | 1.25 | 8.68 | 0.016 |
| IgA Wuhan & IgA Variants | 2.34 | 1 | 5.48 | 0.051 |
| IgG Alinity RBD & IgA Wuhan | 2.91 | 1.12 | 7.6 | 0.029 |

**Table S13.**

Results of Poisson regression comparing infection incidence at 30 follow up days, of the low-baseline and high-baseline response groups using the five primary analysis baseline markers. The model was adjusted to the daily proportion of positive PCR tests to Covid-19, age, occupation, medical center, and time from the third vaccination and subjects as a random effect.

| Marker | Incidence rate ratio | lower .95 | upper .95 | p-value |
|---|---|---|---|---|
| **Three doses** | | | | |
| IgG BioPlex S2 | 1.36 | 0.93 | 2 | 0.118 |
| IgG Alinity RBD | 1.53 | 1.03 | 2.27 | 0.033 |
| IgG Mutants RBD | 1.42 | 0.81 | 2.48 | 0.222 |
| IgA Variants | 1.3 | 0.74 | 2.27 | 0.363 |
| IgA Wuhan | 1.05 | 0.6 | 1.83 | 0.859 |
| **Four doses** | | | | |
| IgG BioPlex S2 | 1.19 | 0.6 | 2.36 | 0.619 |
| IgG Alinity RBD | 1.44 | 0.48 | 4.27 | 0.516 |
| IgG Mutants RBD | 1.57 | 0.56 | 4.46 | 0.393 |
| IgA Variants | 4.53 | 1.22 | 16.82 | 0.024 |
| IgA Wuhan | 2.71 | 0.91 | 8.05 | 0.073 |

**Table S14.**

Results of Poisson regression comparing infection incidence at 60-90 follow up days, of the low-baseline and high-baseline response groups using the five primary analysis baseline markers. The model was adjusted to the daily proportion of positive PCR tests to Covid-19, age, occupation, medical center, and time from the third vaccination and subjects as a random effect.

| Marker | Incidence rate ratio | lower .95 | upper .95 | p-value |
|---|---|---|---|---|
| **Three doses** | | | | |
| IgG BioPlex S2 | 1.46 | 1.06 | 2 | 0.02 |
| IgG Alinity RBD | 1.56 | 1.13 | 2.15 | 0.007 |
| IgG Mutants RBD | 1.49 | 0.93 | 2.38 | 0.096 |
| IgA Variants | 1.38 | 0.87 | 2.18 | 0.173 |
| IgA Wuhan | 1.25 | 0.79 | 1.96 | 0.335 |
| **Four doses** | | | | |
| IgG BioPlex S2 | 1.69 | 1.03 | 2.76 | 0.037 |
| IgG Alinity RBD | 1.67 | 1.02 | 2.74 | 0.04 |
| IgG Mutants RBD | 2.2 | 0.99 | 4.89 | 0.052 |
| IgA Variants | 1.82 | 0.87 | 3.8 | 0.11 |
| IgA Wuhan | 1.88 | 0.95 | 3.69 | 0.069 |

**Table S15.**

Results of Poisson regression comparing infection incidence at 30 follow up days of the low-baseline and high-baseline response groups using all pairwise combinations of the five primary analysis baseline markers. The model was adjusted to the daily proportion of positive PCR tests to Covid-19, age, occupation, medical center, and time from the third vaccination and subjects as a random effect.

| Marker combination | Incidence rate ratio | lower .95 | upper .95 | p-value |
|---|---|---|---|---|
| **Three doses** | | | | |
| IgG Mutants RBD & IgA Variants | 2.92 | 0.75 | 11.27 | 0.121 |
| IgG Alinity RBD & IgA Variants | 2.86 | 1.16 | 7.08 | 0.023 |
| IgG Alinity RBD & IgG Mutants RBD | 1.83 | 0.97 | 3.44 | 0.062 |
| IgG Mutants RBD & IgA Wuhan | 2.64 | 0.69 | 10.06 | 0.155 |
| BioPlex IgG S2 & IgG Alinity RBD | 1.68 | 1.06 | 2.66 | 0.028 |
| BioPlex IgG S2 & IgG Mutants RBD | 1.76 | 0.89 | 3.47 | 0.101 |
| BioPlex IgG S2 & IgA Variants | 2.58 | 1.09 | 6.09 | 0.031 |
| BioPlex IgG S2 & IgA Wuhan | 1.52 | 0.66 | 3.46 | 0.323 |
| IgA Wuhan & IgA Variants | 1.39 | 0.69 | 2.82 | 0.359 |
| IgG Alinity RBD & IgA Wuhan | 1.53 | 0.66 | 3.53 | 0.319 |
| **Four doses** | | | | |
| IgG Mutants RBD & IgA Variants | 5.48 | 0.51 | 59.07 | 0.16 |
| IgG Alinity RBD & IgA Variants | 4.71 | 1.12 | 19.76 | 0.034 |
| IgG Alinity RBD & IgG Mutants RBD | 1.3 | 0.36 | 4.64 | 0.688 |
| IgG Mutants RBD & IgA Wuhan | 3.63 | 0.59 | 22.24 | 0.163 |
| BioPlex IgG S2 & IgG Alinity RBD | 1.43 | 0.58 | 3.54 | 0.443 |
| BioPlex IgG S2 & IgG Mutants RBD | 1.61 | 0.35 | 7.35 | 0.536 |
| BioPlex IgG S2 & IgA Variants | 2.14 | 0.52 | 8.77 | 0.29 |
| BioPlex IgG S2 & IgA Wuhan | 1.89 | 0.51 | 6.99 | 0.339 |
| IgA Wuhan & IgA Variants | 4.49 | 0.92 | 21.78 | 0.063 |
| IgG Alinity RBD & IgA Wuhan | 2.08 | 0.61 | 7.09 | 0.244 |

**Table S16.**

Results of Poisson regression comparing infection incidence at 60-90 follow up days of the low-baseline and high-baseline response groups using all pairwise combinations of the five primary analysis baseline markers. The model was adjusted to the daily proportion of positive PCR tests to Covid-19, age, occupation, medical center, and time from the third vaccination and subjects as a random effect.

| Marker combination | Incidence rate ratio | lower .95 | upper .95 | p-value |
|---|---|---|---|---|
| **Three doses** | | | | |
| IgG Mutants RBD & IgA Variants | 4.66 | 1.29 | 16.87 | 0.019 |
| IgG Alinity RBD & IgA Variants | 2.96 | 1.38 | 6.35 | 0.005 |
| IgG Alinity RBD & IgG Mutants RBD | 1.85 | 1.09 | 3.13 | 0.022 |
| IgG Mutants RBD & IgA Wuhan | 4.5 | 1.28 | 15.87 | 0.019 |
| BioPlex IgG S2 & IgG Alinity RBD | 1.85 | 1.25 | 2.73 | 0.002 |
| BioPlex IgG S2 & IgG Mutants RBD | 1.68 | 0.96 | 2.93 | 0.07 |
| BioPlex IgG S2 & IgA Variants | 2.55 | 1.19 | 5.44 | 0.016 |
| BioPlex IgG S2 & IgA Wuhan | 2.22 | 1.08 | 4.58 | 0.031 |
| IgA Wuhan & IgA Variants | 1.47 | 0.83 | 2.61 | 0.189 |
| IgG Alinity RBD & IgA Wuhan | 1.98 | 0.98 | 4 | 0.058 |
| **Four doses** | | | | |
| IgG Mutants RBD & IgA Variants | 6.87 | 1.26 | 37.38 | 0.026 |
| IgG Alinity RBD & IgA Variants | 3.33 | 1.21 | 9.16 | 0.02 |
| IgG Alinity RBD & IgG Mutants RBD | 2.24 | 0.9 | 5.55 | 0.083 |
| IgG Mutants RBD & IgA Wuhan | 6.45 | 1.4 | 29.6 | 0.017 |
| BioPlex IgG S2 & IgG Alinity RBD | 1.99 | 1.08 | 3.66 | 0.027 |
| BioPlex IgG S2 & IgG Mutants RBD | 2.61 | 0.91 | 7.48 | 0.075 |
| BioPlex IgG S2 & IgA Variants | 2.35 | 0.9 | 6.13 | 0.08 |
| BioPlex IgG S2 & IgA Wuhan | 2.85 | 1.14 | 7.15 | 0.025 |
| IgA Wuhan & IgA Variants | 2.07 | 0.9 | 4.73 | 0.086 |
| IgG Alinity RBD & IgA Wuhan | 2.93 | 1.11 | 7.72 | 0.03 |
